# Common and rare variant analyses reveal novel genetic factors underlying Idiopathic Pulmonary Fibrosis and shared aetiology with COVID-19

**DOI:** 10.1101/2024.08.16.24312138

**Authors:** Athanasios Kousathanas, Christopher A. Odhams, James Cook, Yao Hu, Stephan Klee, A. Mesut Erzurumluoglu, Fidel Ramirez, Christoph Mayr, Dennis Schäfer, Lara Beck, Ingrid Christ, Taekyu Lee, James Christopher Tarr, Steven W. Fesik, Boehringer Ingelheim – Global Computational Biology and Digital Sciences, James Duboff, Frederik Wirtz-Peitz, Loukas Moutsianas, Matthew A. Brown, Jan Kriegl, Goerge Okafo, Jan N Jensen, Matthew J. Thomas, Zhihao Ding

## Abstract

Idiopathic pulmonary fibrosis (IPF) is a progressive and debilitating respiratory disease with limited therapeutic options. We carried out genome-wide association (GWAS), post-GWAS and rare variant analyses utilising the whole genome sequencing data (WGS) from the 100,000 Genomes Project (100kGP) of a cohort of IPF participants (*n*=586) to identify novel associations and potential drug targets. Meta-analysis combining 100kGP and published GWASs of IPF (total 11,746 cases and 1,416,493 controls) identified a novel association signal at the 1q21.2 locus (rs16837903, OR[95%CI]=0.88[0.85, 0.92], *P*=9.54×10^−9^) which was also successfully replicated with independent data and linked to the probable effector gene *MCL1. MCL1* showed increased expression levels in IPF patients versus controls in alveolar epithelial type I cells. Despite its known antiapoptotic role, inhibition of *MCL1 in vitro* did not selectively deplete senescent cells, hinting at the complexity involved in targeting *MCL1*. Rare variant burden analysis identified *ANGPTL7*, a secreted glycoprotein involved in the regulation of angiogenesis, as a novel IPF candidate gene (OR[95%CI]=28.79 [8.51-97.43], *P*=6.73×10^−8^). Transcriptome-wide association analysis (TWAS) revealed that overexpression of cell cycle regulator *SERTAD2* and nuclear importer *TPNO3* were associated with increased IPF risk. We also investigated shared genetic mechanisms between IPF with severe COVID-19 and expanded the list of shared genetic loci with three novel colocalised signals at 1q21.2, 6p24.3 and 16p13.3 with probable effector genes *MCL1*, *DSP and RHBDF1*, implicating regulation of apoptosis, cell adhesion and epidermal growth factor signalling, respectively. By leveraging the genetic correlation between IPF and severe COVID-19 (*r_g_*[95% CI]) = 0.39 [0.25-0.53]) through multi-trait meta-analysis, we further identified and replicated an additional novel candidate IPF signal at 2p16.1 with probable effector gene *BCL11A*, a regulator of haematopoiesis and lymphocyte development. Based on prioritized genes across analyses, we propose mechanisms mediating IPF disease risk and shared mechanisms between IPF and severe COVID-19, thereby expanding the potential for developing common treatments.

## Introduction

Idiopathic pulmonary fibrosis (IPF) is a debilitating lung disease characterised by irreversible loss of lung function. IPF has a poor prognosis and limited treatment options that only slow disease progression rather than being curative^1,2^, presenting a major unmet therapeutic need.

Due to the mechanisms of the disease being largely unknown, hypothesis-free genetic studies are particularly attractive for identifying novel genes and pathways relevant for IPF pathology and for identifying potential new treatment targets. Genome-wide association studies (GWAS) have been successful in identifying several genetic variants associated with IPF, implicating pathways involved in host defence, telomere maintenance, mTOR signalling, and cell-cell adhesion^3^. As IPF is a rare disease, GWAS discovery for IPF has been powered by consortia that aggregate Biobank data around the world through meta-analysis^3–5^. The latest and best-powered effort was led by the Global Biobank Meta-analysis initiative (GBMI) that assembled multi-ancestry data from 13 Biobanks and from the International IPF Genetics Consortium, to identify 25 independent genetic associations with IPF of which three were replicated novel associations^5^.

Most published IPF genetic studies have employed SNP microarrays for genotyping. However, this has limited the investigation of rare pathogenic variants contributing to IPF susceptibility. More recently, whole exome and whole genome sequencing (WES/WGS) rare variant studies have added evidence for rare variant effects on telomere-related genes (*TERT*, *RTEL1*)^6,7^, mRNA stability and degradation (*PARN*) and spindle assembly during cell division (*SPDL1* and *KIF15*)^8,9^.

Infectious diseases with a strong respiratory component have been linked to the pathogenesis of IPF^10^ and the recent COVID-19 pandemic prompted studies on potential genetic overlaps between IPF and severe COVID-19^5,11,12^ enabled by the wealth of data that has been generated for the latter^13–15^. Exploring the connection between IPF and severe COVID-19 could provide additional therapeutic concepts and potentially increase statistical power for genetic target discovery in IPF.

The 100,000 Genomes Project (100kGP) is one of the world’s largest rare disease projects, in which undiagnosed patients with suspected monogenic diseases were recruited and screened by WGS to aid in their diagnosis. 100kGP participants include people affected with familial forms of IPF, as well both prevalent and incident sporadic cases, the latter identified by data linkage with electronic health records (EHRS).

In this study, we set out to perform the largest meta-analysis of IPF by leveraging the 100kGP resource and GBMI GWAS summary statistics to further enhance power of common variant analysis^1^ as well as to search for associations in the rare variant space enabled by the 100kGP WGS data. We also further explored the genetic overlap of IPF and severe COVID-19 and leveraged the correlation of IPF and severe COVID-19 to amplify statistical power for GWAS discovery.

## Results

### The 100,000 Genomes Project (100kGP) IPF cohort

We defined an Idiopathic Pulmonary Fibrosis (IPF) case cohort of (*n*=586) individuals from participants of the 100,000 Genomes Project (100kGP). IPF cases comprised of individuals that were specifically recruited into the 100kGP for familial pulmonary fibrosis (n=147) plus ‘incidental’ cases of IPF identified in the 100kGP cohort through analysis of linked EHRs (n=439, Supplementary Tables 1-4). We used a set of 100kGP participants (n=52,083) as controls after excluding individuals with relevant respiratory EHR indications (Supplementary Table 2) and examining matching of demographic characteristics such as age, sex and ancestry with cases to inform covariate selection downstream (Supplementary Tables 5-6 and Supplementary Figures 1-2, Methods). We kept 100kGP individuals with European genetically predicted ancestry and with WGS data that had passed sample quality control (Methods).

### IPF GWAS analysis and meta-analysis

#### 100kGP GWAS

We performed GWAS analysis for the 100kGP IPF cohort with *N_cases_*=586 and *N_controls_*=52,083 on 11,646,047 common variants (MAF>0.5%) that yielded one genome-wide significant (*P*-value<5×10^−8^) signal at locus 11p15.5 with top variant chr11:1219991_G_T (rs35705950), *P*-value = 2.09×10^−25^, OR [95%CI] = 2.94 [2.40, 3.60] (Supplementary Figure 3). This is the well-known IPF-associated common variant in the promoter region of *MUC5B*^16^.

#### Meta-analysis

We then meta-analysed the 100kGP IPF GWAS results with the Partanen et al. (2022)^5^ GWAS summaries (Supplementary Table 7) with inverse variance weighted meta-analysis for 10,644,973 variants (Figure 1). We used GCTA-COJO^17^ to identify 34 conditionally independent signals reaching genome-wide significance (*P*-value<5×10^−8^) across 23 loci (Supplementary Table 8; Methods). 19 loci harboured a single independent signal each, and the remaining four loci at 5p15.33, 11p15.5, 15q15.1 and 19p13.3, contained more than two independent signals each (Supplementary Table 8).

**Figure 1.**
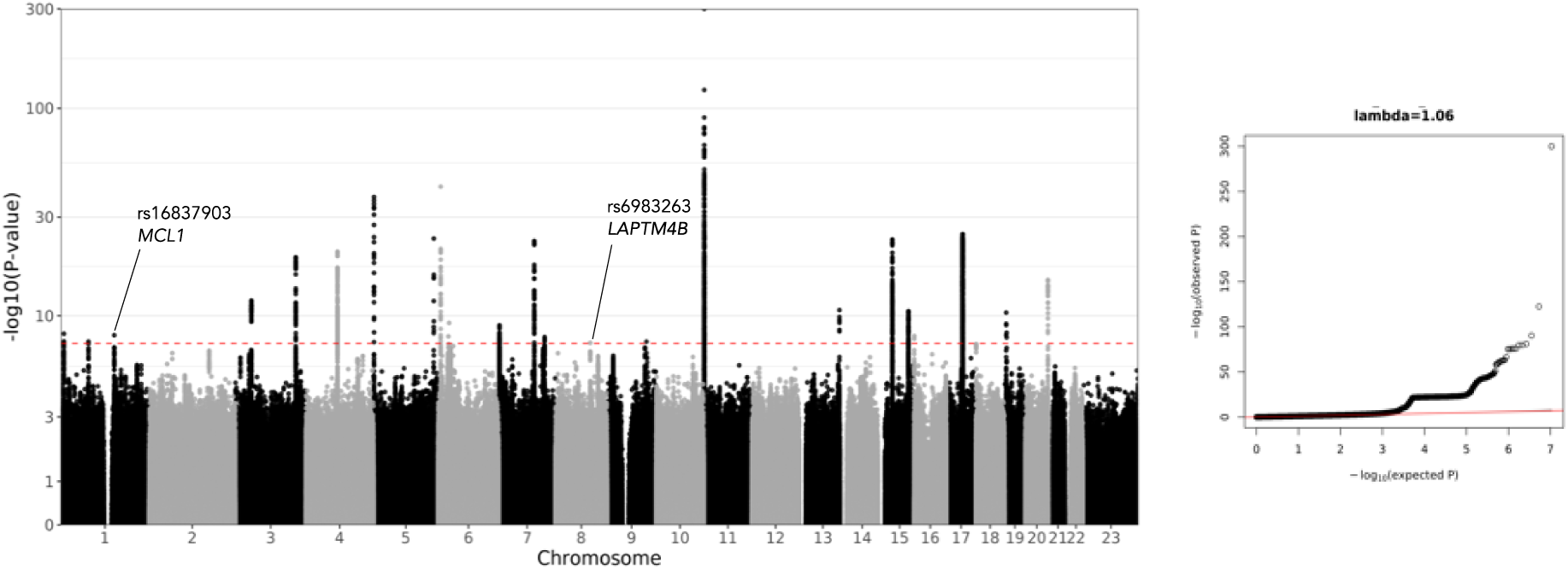
Manhattan and Q-Q plot for the IPF meta-analysis of Partanen et al. 2022 and 100kGP GWAS.

We mapped lead variant signals to genes by annotating variant effects with Variant Effect Predictor (VEP) and also mapped the nearest gene for each lead variant (Table 1; Supplementary Table 8). More than half of the identified variants were in introns, while we identified two variants with missense consequences and six with regulatory consequences (Supplementary Table 8). Fine-mapping with SusieR^18^ enabled identification of credible sets (CS) that enabled prioritisation of additional variants by identifying those that had the worst consequence across the CS (Supplementary Table 8).

**Table 1:** Novel meta-analysis independent signals reaching genome-wide significance. Loci are displayed as cytogenetic band locations of the variants. For each variant, two IDs are shown: The variant ID, consisting of Chromosome, position in hg38 coordinates, reference and alternate hg38 alleles (chr:pos_ref_alt), and the respective rsID. AF EUR corresponds to the allele frequency of the variants in a set of N=44,590 unrelated European individuals from the 100kGP that was used also as the LD reference for the conditional analysis. Odds ratios (OR) are calculated for the alternate allele matching the variant ID specification. N_var_ clump corresponds to the number of variants included in the LD clump to which the displayed variant belongs. Full meta-analysis discovery table is Supplementary Table 8. Associations with a significant P-value that is less than a Bonferroni corrected threshold of 0.05/2=0.025 and with consistent odds ratio between discovery and replication datasets are annotated with asterisk (*) in replication P-value column. For each lead variant, VEP v.105-assigned most severe consequence across transcripts and the impacted gene is shown, with the nearest gene in parentheses when different.

| Locus | Lead variant ID (rsID) | AF EUR | OR (95%CI) | $P$-value | Replication OR (95%CI) | Replication $P$-value | $N_{var}$ clump | VEP Consequence | VEP Gene (Nearest) |
| --- | --- | --- | --- | --- | --- | --- | --- | --- | --- |
| 1q21.2 | chr1:150579566_G_A (rs16837903) | 0.15 | 0.88 (0.85, 0.92) | 9.54x10<sup>-9</sup> | 0.82 (0.72, 0.95) | 0.0057* | 239 | 5' UTR | *MCL1* |
| 8q22.1 | chr8:97822998_T_C (rs6983263) | 0.48 | 0.92 (0.9, 0.95) | 4.47x10<sup>-8</sup> | 0.91 (0.8, 1.03) | 0.124 | 201 | Intron | *LAPTM4B* (*MATN2*) |

Two of the identified associations were novel ^55^: (1) Signal at locus 1q21.2 with top variant chr1:150579566_G_A; rs16837903; OR (95%CI) = 0.88 [0.85, 0.92]; *P*-value=9.54×10^−9^ which is a 5’ UTR regulatory variant of *MCL1* (Table 1; Supplementary Figure 4). (2) Signal at locus 8q22.1 with top variant chr8:97822998_T_C; rs6983263; OR (95%CI) = 0.92 (0.9, 0.95); *P*-value=4.47×10^−8^ which is an intronic variant of *LAPTM4B* (Table 1; Supplementary Figure 5).

#### Replication of IPF novel signals

To replicate the two novel signals identified in the meta-analysis (*Table 1*) we generated an independent IPF dataset through mathematical subtraction from another, partially overlapping, IPF GWAS dataset^4^ (Methods). Both novel variant signals had consistent direction of effects in the replication dataset, but only rs16837903 had a significant replication *P*-value (*P*<0.025; *Table 1*).

### IPF aggregate rare variant burden testing (AVT) analysis

We expanded the analysis to rare variants that could potentially underlie additional associations with IPF by performing aggregate rare variant burden testing (AVT) analysis for the 100kGP IPF cohort with *N_cases_*=569 and *N_controls_*=50,847 for bi-allelic variants with MAF<0.5% (Supplementary Table 10). We ran AVT analyses for four variant masks: loss of function, ultra-rare damaging, rare damaging and flexible damaging (Supplementary Table 11). These analyses identified two genes, *ANGPTL7* and *TERT*, that achieved study-wide significance (*P*-value < 6.81×10^−7^) in at least one mask for the composite SKAT-O test (Table 2, Supplementary Figure 6). *ANGPTL7* was significant in the loss-of-function mask (SKAT-O *P*-value = 5.64×10^−7^), while *TERT* was significant in all but the loss of function mask (Table 2).

**Table 2:** Association results across masks for significant genes in the burden analysis. The total number of variants included in each test is shown (# Variants) and the number of variants included in the collapsed ultra-rare variant (# Ultra-rare). Study-wide significant associations for the SKAT-O test correcting for the total number of tests across all four masks (P-value < 6.81×10^7^) are annotated with asterisk (*). Dashes correspond to entries where summary statistics were not calculated due to all variants being ultra rare.

| Gene | Mask | # Variants | # Ultra-rare | P-value SKAT-O | P-value Burden | BETA burden (SE) |
| --- | --- | --- | --- | --- | --- | --- |
| **ANGPTL7** | LOF | 5 | 4 | $5.64 \times 10^{-7*}$ | $1.09 \times 10^{-6}$ | 0.128 (0.0262) |
| | RD | 55 | 53 | 1 | $9.07 \times 10^{-1}$ | -0.00464 (0.0395) |
| | URD | 37 | 37 | $5.39 \times 10^{-1}$ | - | - |
| | FD | 74 | 65 | $2.56 \times 10^{-6}$ | $5.84 \times 10^{-2}$ | 0.029 (0.0153) |
| **TERT** | LOF | 7 | 7 | $7.78 \times 10^{-2}$ | - | - |
| | RD | 58 | 58 | $7.05 \times 10^{-8*}$ | - | - |
| | URD | 49 | 49 | $3.01 \times 10^{-8*}$ | - | - |
| | FD | 102 | 100 | $2.17 \times 10^{-10*}$ | $1.32 \times 10^{-9}$ | 0.114 (0.0187) |

LOF: loss-of-function, RD: rare damaging, URD: ultra-rare damaging, FD: flexible damaging.

Rare deleterious genetic variants in *TERT* have been previously robustly associated with IPF^19^, whereas *ANGPTL7* represents a novel association signal. We investigated further the individual variant statistics contributing to the signal for *ANGPTL7*, and identified a single variant chr1:11193631_C_T (rs143435072, HGVSp p.Arg177Ter) as the main driver of the signal with *P*-value=6.73×10^−8^ and OR[95%CI]=28.79 [8.51-97.43] (Supplementary Tables 12-13). Allele frequency for rs143435072 was 7.91x-10^−3^ in cases and 3.05×10^−4^ in controls, the latter being similar to allele frequency for non-Finnish Europeans (3.24×10^−4^) in GnomAD v3.1.2, indicating that control composition is unlikely to underlie this signal.

### IPF Transcriptome-wide association study analysis (TWAS)

To assess how genetically determined variation in gene expression might impact IPF disease susceptibility, we conducted a transcriptome-wide association study (TWAS) with S-prediXcan^20^ utilizing gene expression data (GTEx v8) from lung tissue and leveraging the updated and better-powered IPF summaries from this study compared to previous TWAS analyses^21,22^. We tested a total of 14,528 genes and identified 21 genes with a significant *P*-value at Bonferroni adjusted threshold (P<3.44×10^−6^; Figure 2). As TWAS signals can be driven by spurious pleiotropy, we also ran colocalisation^23^ analysis to test for shared causal variants between lung gene expression and IPF susceptibility. Seven genes successfully colocalised, with *TNPO3* and *SERTAD2* being previously unreported^21,22^ TWAS signals (Supplementary Table 14).

**Figure 2.**
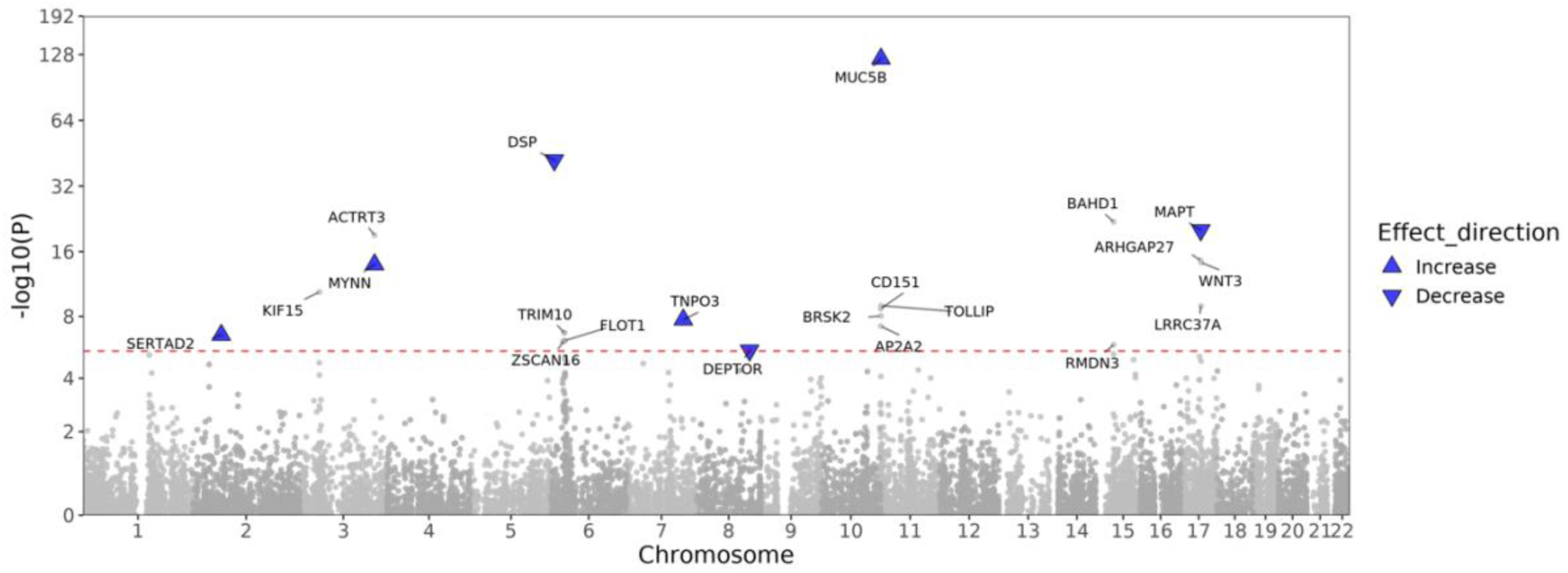
Gene-wide Manhattan plot from a Transcriptome-wide association study (TWAS) for IPF using S-prediXcan and GTEX v8 expression data from lung tissue. Annotated genes have significant P-values at the Bonferroni adjusted significance threshold of 0.05/14,528=3.44×10^−6^ (red dashed line). Genes highlighted in blue successfully colocalised with coloc in lung tissue (Methods). Effect direction corresponds to the predicted change in gene expression and is obtained from the gene-level z-score sign.

#### IPF and COVID-19

The availability of updated COVID-19 GWAS summary statistics compared to previous studies^5^ enabled us to revisit the genetic overlap of IPF and severe COVID-19 by conducting colocalisation analysis. We also re-estimated and leveraged the genetic correlation between the traits to amplify statistical power to discover IPF associations by performing multi-trait meta-analysis (MTAG)^24^. For these analyses we used the IPF meta-analysis statistics produced in this study and the latest meta-analysis summaries (Freeze 7) from the COVID-19 Host Genetics Initiative (HGI), for the Hospitalised covid vs. population phenotype (B2_ALL_leave_23andme), as this has produced the best balance between a carefully curated phenotype for severity and statistical power^15,25^(Supplementary Table 15).

We tested for colocalisation between IPF and severe COVID-19 for a total of 23 loci significantly associated with IPF, and obtained significant colocalization evidence for six loci (Table 3, Supplementary Figure 7). Three signals at loci 11p15.5, 13q34 and 19p13.3 with lead variants with probable effector genes *MUC5B*, *ATP11A* and *DDP9* reproduce signals previously reported^12^, while an additional three signals at 1q21.2, 6p24.3 and 16p13.3 with probable effector genes *MCL1*, *DSP and RHBDF1*, represent novel colocalization signals (Table 3).

**Table 3.**
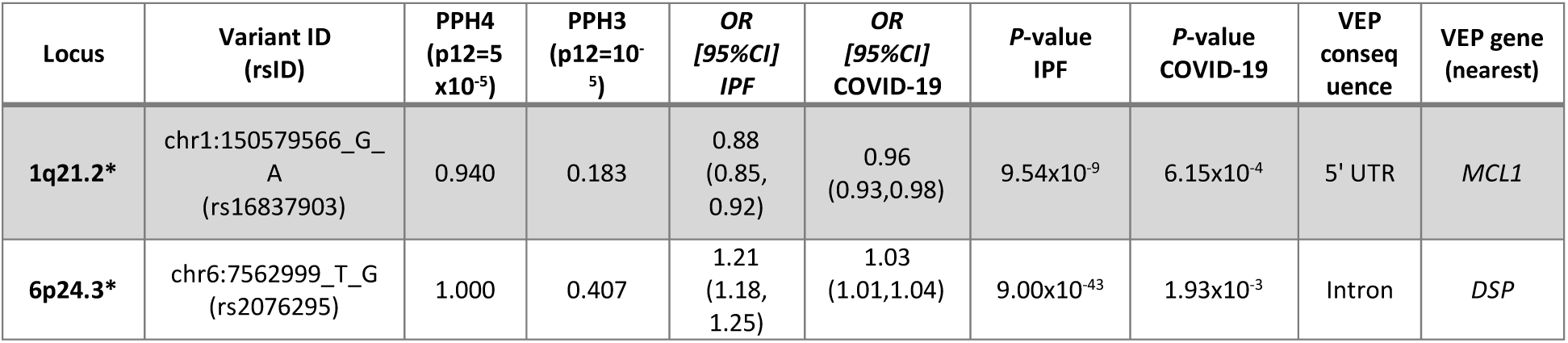
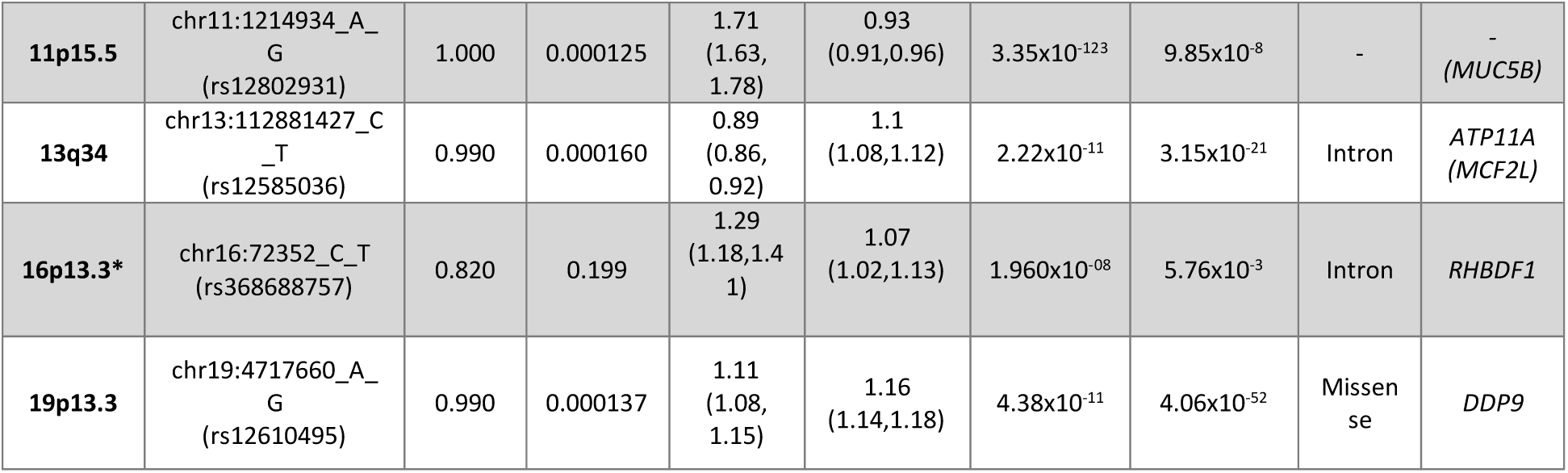
Colocalisation results between IPF and Severe COVID-19. Posterior probabilities PPH4 and PPH3 which were used to determine significant colocalisation. Novel significant colocalised loci are marked in with an asterisk(*) within the Locus column. The variant with the highest PPH4 in each locus is shown. The variant ID consists of Chromosome, position in hg38 coordinates and reference and alternate hg38 alleles (chr:pos_ref_alt). Odds ratios and P-values for IPF and severe COVID-19 GWAS, VEP v.105 consequences for the variants and affected genes are shown. Nearest gene is displayed in parentheses if it is different than the VEP annotated gene.

We then estimated the genetic correlation between IPF and severe COVID-19 (*r_g_*[95% CI]) as 0.39 [0.25-0.53] (Supplementary Table 16), which is consistent with previous estimates^5^, albeit with tighter confidence intervals.

To amplify statistical power for discovery of new genetic associations with IPF we leveraged the estimated genetic correlation between IPF and severe COVID-19 by performing multi-trait meta-analysis between the two traits. MTAG analysis provided individual trait summary statistics for both IPF and severe COVID-19 with signals amplified or attenuated by the estimated genetic correlation between the traits (Figure 3).

**Figure 3.**
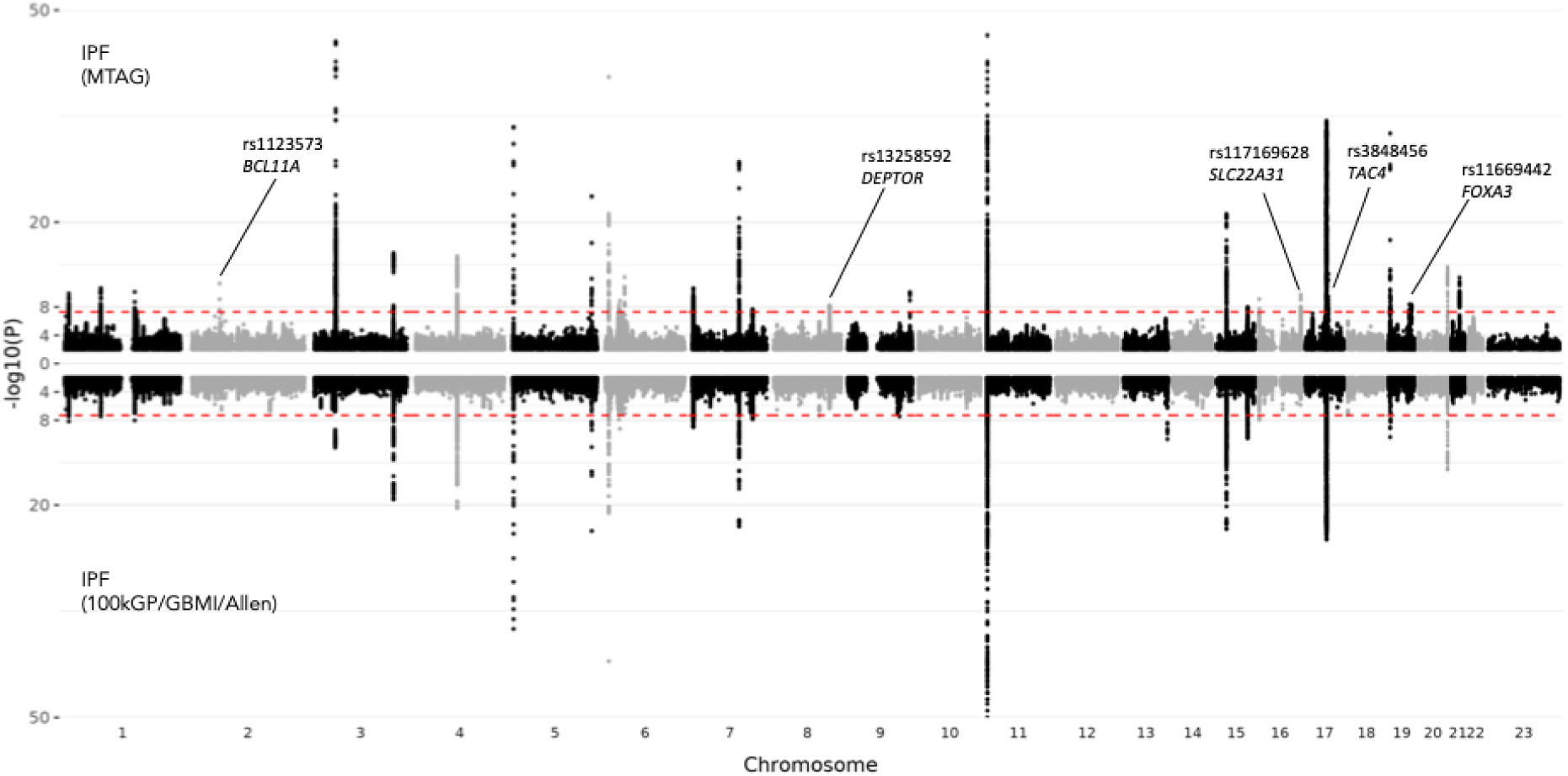
MTAG results for IPF (top) compared with the IPF base meta-analysis (bottom). Red dashed line corresponds to P-value < 5×10^−8^. P-values below 10^−50^, are set to 10^−50^.

With MTAG, we identified five novel IPF-associated loci at 2p16.1, 8q24.12, 16q24.3, 17q21.33, 19q13.32 and mapped VEP consequences for lead variants to *BCL11A*, *DEPTOR*, *SLC22A31*, a regulatory region, and *FOXA3*, respectively (Table 4). We used the independent 2-way Allen et al. 2019 IPF meta-analysis to check replication evidence for these signals. The locus signal at 2p16.1, chr2:60480453_A_G (rs1123573), an intronic variant for *BCL11A*, successfully replicated (Table 4).

**Table 4.** Novel signals from clumping analysis of IPF MTAG results. The P-value for each clump lead variant was required to be genome-wide significant in the MTAG analysis (P<5×10^−8^) and less than 7.04×10^−4^ in the discovery IPF meta-analysis (Bonferroni corrected P-value threshold for the 71 MTAG-significant signals; Methods). Associations with replication P-value less than Bonferroni corrected threshold of 0.05/5=0.01 and with consistent odds ratio between discovery and replication datasets are annotated with an asterisk (*). For each lead variant mapped VEP v.105 consequences and genes are shown. Nearest gene is displayed in parentheses if it is different than the VEP annotated gene.

| Locus | Clump Lead Variant (rsID) | Discovery IPF P-value (COVID) | Discovery IPF OR [95% CI] | MTAG P-value | MTAG IPF OR [95% CI] | Replication OR (95%CI) | Replication P-value | VEP Consequence | VEP gene (nearest) |
| --- | --- | --- | --- | --- | --- | --- | --- | --- | --- |
| **2p16.1** | chr2:60480453_A_G (rs1123573) | $2.31 \times 10^{-5}$<br>($1.32 \times 10^{-14}$) | 0.94<br>(0.91, 0.97) | $4.51 \times 10^{-12}$ | 0.94<br>(0.92, 0.96) | 0.818 | $7.31 \times 10^{-4}$ * | intron | *BCL11A* |
| **8q24.12** | chr8:119900189_G_A (rs13258592) | $2.01 \times 10^{-6}$<br>($1.36 \times 10^{-4}$) | 1.07<br>(1.04, 1.1) | $6.81 \times 10^{-9}$ | 1.06<br>(1.04, 1.07) | 0.908<br>(0.803, 1.03) | 0.124 | intron | *DEPTOR* |
| **16q24.3** | chr16:89196249_G_A (rs117169628) | $5.81 \times 10^{-4}$<br>($1.37 \times 10^{-14}$) | 1.08<br>(1.03, 1.12) | $1.74 \times 10^{-10}$ | 1.06<br>(1.04, 1.08) | 1.15<br>(1.02, 1.31) | 0.0243 | missense | *SLC22A31* |
| **17q21.33** | chr17:49863260_C_A (rs3848456) | $1.25 \times 10^{-4}$<br>($2.56 \times 10^{-21}$) | 1.14<br>(1.06, 1.21) | $2.08 \times 10^{-13}$ | 1.07<br>(1.05, 1.09) | 1.1<br>(0.833, 1.45) | 0.4975 | regulatory region | -<br>(*TAC4*) |
| **19q13.32** | chr19:45871299_T_C (rs11669442) | $3.07 \times 10^{-5}$<br>($1.09 \times 10^{-7}$) | 1.07<br>(1.03, 1.1) | $4.31 \times 10^{-9}$ | 1.05<br>(1.04, 1.07) | 1.07<br>(0.973, 1.18) | 0.160 | intron | *FOXA3* |

#### Functional follow-up of *MCL1*

We further investigated the potential disease mechanism for *MCL1* (the candidate effector gene at the novel IPF locus) by evaluating the expression of *MCL1* in single-cell expression data from single-cell suspensions generated from lung tissue from IPF patients and non-fibrotic controls (GSE135893^26^). We observed that *MCL1* expression was increased in alveolar epithelial type I cells as well as in KRT5-/KRT17+ cells (aberrant basaloid cells) in IPF cases compared to controls (Supplementary Figure 8). We also found that the expression pattern of *MCL1* was similar tissue-wise with the expression of *CDKN1A*, the gene coding for p21 (Supplementary Figure 8), which is a marker of senescence^27^. The occurrence of increased levels of senescence is well described for IPF^28^. Moreover, it has previously been shown that alveolar epithelial cells exhibit an increased level of senescence and contribute to disease progression^29,30^. Considering MCL1 is an anti-apoptotic member of the BCL2 protein family, we hypothesized that inhibiting MCL1 in senescent alveolar epithelial cells will selectively deplete these cells through apoptosis.

We therefore induced cellular senescence in primary human lung epithelial cells enriched for HT2-280-positive cells (marker of ATII cells) using bleomycin (Figure 4). We observed an increase of *CDKN1A* gene expression over time along with an increased expression of p21 (Figure 4 A-B), confirming successful induction of senescence. We then analysed the expression of MCL1 as well as the complexes of MCL1 with BIM and BAK, which are important pro-apoptotic proteins of the BCL2 protein family. Binding of MCL1 to BIM and BAK can suppress pro-apoptotic functions, and conversely, releasing them from the complex can induce these pro-apoptotic functions. However, we did not observe any increased expression of MCL1, and only small increases in the MCL1:BIM and MCL1:BAK complexes (Figure 4 C-E). We additionally tested more senescence- and fibrosis-associated stimuli, but none of these stimuli increased the expression of MCL1 significantly (data not shown).

**Figure 4.**
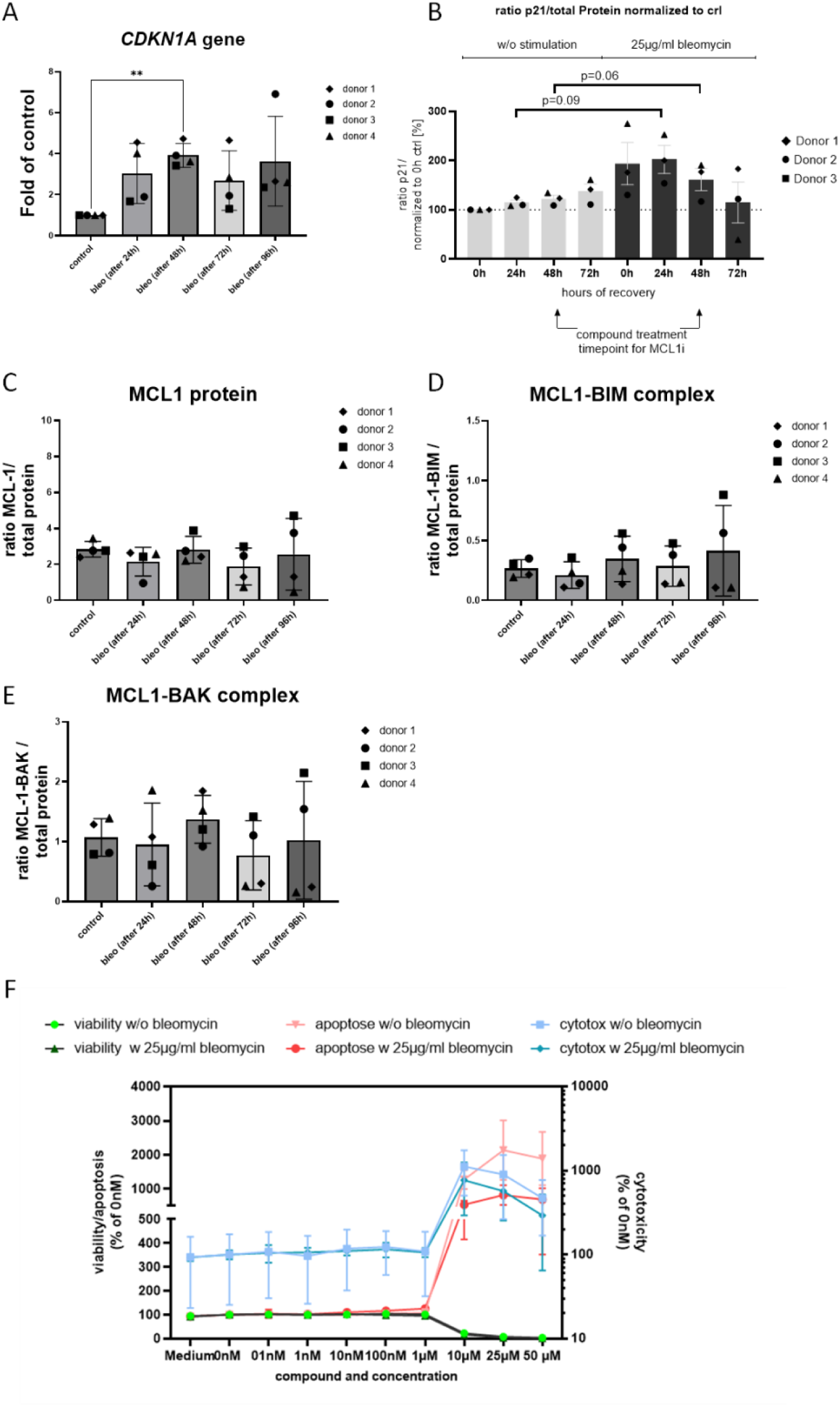
Induction of senescence in primary lung epithelial cells enriched for HT2-280 and subsequent testing of an anti-MCL1 inhibitor. Cells have been treated with bleomycin for indicated timepoints and (A) CDKN1A gene expression, (B) p21 protein staining, (C) MCL1 protein, (D) MCL1:BIM complex or (E) MCL1:BAK complex was assessed. (F) Cells were treated with bleomycin for 24h, then rested for 48h and subsequently treated with an MCL1 antagonist for 24h. The effect of MCL1 antagonism on viability, apoptosis (y-axis left) and cytotoxicity (y-axis right) in healthy (w/o bleomycin) and senescent cells (w/ bleomycin) were measured. (N=3-4 of individual donor cells). Asterisks highlight significant P-values where shown, * P-value < 0.05, ** P-value < 0.01.

Although we could not detect an increase in MCL1 expression, we tested if we nevertheless could specifically deplete senescent alveolar epithelial cells *in vitro*, sparing healthy cells. We did not observe a difference of effect when comparing senescent and healthy cells, as both showed decreases in viability and increases in apoptosis and/or cytotoxicity at similar proportions (Figure 4F).

Taken together, we observed increased *MCL1* expression in IPF patients using single cell expression data, but inhibition of MCL1 failed to specifically deplete senescent lung epithelial cells.

## Discussion

In this study we performed the largest meta-analysis of IPF to date by integrating GWAS results from the 100,000 Genomes Project (100kGP) IPF cohort with previous meta-analysis summary statistics to increase GWAS discovery power and we also leveraged WGS data for rare variant analysis of this cohort. The increased statistical power of common variant meta-analysis allowed us to discover and replicate a novel association signal at 1q21.2 with probable effector gene *MCL1*. Rare variant burden analysis revealed a novel candidate gene association for *ANGPTL7* and TWAS analysis prioritised additional genes mediating risk for IPF through effects on expression in lung (*SERTAD2* and *TNPO3*). We also updated the genetic correlation estimate and colocalised signals with severe COVID-19, and for the first time we leveraged that correlation through multi-trait meta-analysis to discover a novel and robustly replicated association signal (at 2p16.1/*BCL11A*). Following up on the novel GWAS signal from this study, we also investigated potential mechanisms linking probable effector gene *MCL1* expression to IPF via *in silico* and *in vivo* analyses with a focus on the gene’s known anti-apoptotic properties and exploration of this gene as a therapeutic target.

### *MCL1* and potential as a therapeutic target

The lead variant from the meta-analysis GWAS, rs16837903, is in the 5’UTR of myeloid leukaemia 1 (*MCL1*) gene. MCL1 is an anti-apoptotic member of the BCL-2 family, which are regulators of apoptosis^31^ and are involved in various (patho-)physiological events, including development, wound repair, and cancer. IPF is a disease characterized by uncontrolled wound repair in the accumulation of senescent cells, both of epithelial and mesenchymal origin^32^. In a healthy environment, senescent cells initially contribute to tissue remodeling and repair and are subsequently cleared by NK cells or macrophages^32^. However, this process is impaired in IPF, and it has previously been shown that several IPF-relevant mechanisms can lead to an increase in senescence, including TGF-β signalling, telomere attrition, and mitochondrial dysfunction^33^. Members of the BCL2 protein family, in particular the anti-apoptotic members BCL2 and BCL-XL, have been implicated to block apoptosis in these cells and thereby contribute to IPF pathogenesis^34,35^. General ablation of senescent cells ^36^ but also specific targeting of BCL2 and BCL-XL reduced the levels of senescence and fibrotic burden both in *in vitro* cultures and *in vivo* lung fibrosis models^34^. However, targeting BCL2 and BCL-XL in IPF might not be feasible, as current pharmacological interventions are associated with a broad range of side effects and further investigations are needed to address this^37^. The identification of *MCL1* as a probable effector gene offered a potential new avenue of targeting senescent cells in IPF, which was further supported by the upregulated *MCL1* level in epithelial subsets based on single cell sequencing data. However, we did not detect an increase in *MCL1* expression in induced senescent primary lung epithelial cells from different donors. Previous studies have shown that increased expression of MCL1, and other BCL2 proteins like BCL2 or BCL-XL, is an important prerequisite to selectively deplete cancer cells^38,39^. Even though *MCL1* was not significantly upregulated in senescent cells, we found that pro-apoptotic members (BIM and BAK and their complexes with MCL1) were increased in the senescent cells, indicating MCL1 inhibition may have put an additional apoptotic pressure on senescent primary lung epithelial cells. We tested the effects of an MCL1 inhibitor on its ability to selectively deplete senescent primary lung epithelial cells through inducing apoptosis. However, we did not see an earlier induction of apoptosis in these cells, which was comparable to healthy primary lung epithelial cells along with effects on viability and cytotoxicity. Alongside the HT2-280-enriched cells, we have also tested small airway epithelial cells (data not shown) and observed similar results for both. We therefore could not achieve a clear selective depletion. In IPF, not only epithelial cells are shown to have an increased senescence phenotype, but also immune and mesenchymal cells^40,41^. Therefore, while our studies indicate that epithelial cells might not be the prime target for an anti-MCL1 strategy to induce senolytic effects and improve the outcome for IPF patients, targeting other cell types might have beneficial effects. Further studies on *MCL1* will be required to address these questions.

### Rare variants and *ANGPTL7*

Rare variant analysis identified *ANGPTL7* loss of function variant rs143435072 as a strong risk-increasing risk factor for IPF. rs143435072 has previously been associated with lower intraocular pressure and protection against glaucoma ^42^. ANGPTL7 belongs to the functionally heterogeneous family of angiopoietin-like proteins (ANGPTLs)^43–45^. Apart from glaucoma^42,46^, it has also been implicated in corneal angiogenesis^47^, and tumour growth^48^. In the lung, *ANGPTL7* is specifically expressed in type-1 alveolar epithelial cells (AT1)^49^. We performed a systematic review of the expression of *ANGPTL7* in the lung using IPF Cell Atlas^49^ to develop hypotheses for a potential role of *ANGPTL7* on IPF (Supplementary Material, Supplementary Figures 9-11). In brief, we hypothesise that ANGPTL7 may contribute to IPF by influencing interstitial ECM formation or by inhibiting alveolar angiogenesis. A critical next step is to replicate the *ANGPTL7* rare variant signal identified here in other independent cohorts. Further investigations using functional assays and mouse models could then help to elucidate the precise involvement of *ANGPTL7* in the lung and IPF pathogenesis.

### Gene expression effects of *SERTAD2* and *TNPO3*

Transcriptome wide association analysis revealed that overexpression of *SERTAD2* and *TNPO3* in lung is associated with increased IPF risk. *SERTAD2*, also known as TRIP-Br2, is a cell cycle transcriptional co-regulator that has been linked to regulation of lipolysis, thermogenesis and oxidative metabolism^50^. *SERTAD2* has been shown to promote oncogenesis in nude mice, is overexpressed in multiple human tumors^51^ and is activated by DNA repair protein *REV1* in a Rad18-dependent manner to promote lung tumorigenesis^52^. These previous observations suggest that *SERTAD2* overexpression might mediate risk for IPF through dysregulation of DNA repair. In turn, *TNPO3* (Transportin 3) is a nuclear import receptor for serine/arginine-rich (SR) proteins. Interestingly, previous studies have shown that mutations in *TNPO3* can cause limb girdle muscular dystrophy^53^, in some cases through interference in the morphology and function of the myofibrillar network^54^. Interestingly, GWAS identified variants within the *TNPO3-IRF5 locus* have been previously associated with systemic sclerosis^55,56^, another fibrotic disease characterized by the excessive accumulation of collagen and other extracellular matrix proteins in the skin and various internal organs. Follow-up work with colocalisation analysis between IPF, systemic sclerosis and cis-eQTLs for this locus could help test whether causal variants are shared and potentially underlie a common disease mechanism.

### IPF and COVID-19: colocalisation

Several previous studies have set out to identify shared genetic signals between IPF and severe COVID-19 phenotypes to investigate potential shared aetiology and mechanisms between the two diseases^5,11,12^. Following similar colocalisation analysis and using updated IPF and severe COVID-19 summaries we identified novel shared signals at 1q21.2, 6p24.3 and 16p13.3 with *MCL1*, *DSP* and *RHBDF1* as probable effector genes.

As discussed above, dysregulation of *MCL1* could contribute to IPF by enabling survival of epithelial cells exacerbating fibrosis. *MCL1* could follow an analogous mechanism in contributing to severe COVID-19 through dysregulation of apoptosis for SARS-CoV-2 infected cells. Remarkably, a recent study showed that SARS-CoV-2 suppresses apoptosis in cultured cells, human organoids and mice by interacting with *MCL1*^57^, a mechanism also known as *virus-induced senescence*^58^. *DSP* is a key component of desmosomes, structures involved in cell-cell adhesion in epithelial tissues, including the lungs and is already established as a risk factor for IPF^59^. A recent study found that acute COVID-19 patients had increased levels of desmosomal Desmoglein-2 (DSG2) protein antibodies in serum contributing to an autoimmune mechanism potentially by exacerbating cardiac related complications^60^. *RHBDF1* encodes a intramembrane serine protease involved in the regulation of EGFR (Epidermal Growth Factor Receptor) signalling, which is critical for cell proliferation, survival, and wound healing^61^. Activation of EGFR through binding of either EGF or EGF-like ligands, such as TGF- α, triggers downstream signalling pathways (e.g., JAK/STAT) that promote the progression of cancer and virus infection and targeting the EGFR signalling pathway has been proposed as a potential therapeutic option for severe COVID-19^62^.

### IPF and COVID-19: multi-trait analysis

In this study for the first time, we also leveraged the genetic correlation between IPF and severe COVID-19 for GWAS discovery through multi-trait meta-analysis. The moderate genetic correlation between these traits (*r_g_*=0.39) suggests that multi-trait meta-analysis is expected *a priori* to have limited benefits in terms of amplifying statistical signal^63^. However, our analysis identified five novel IPF locus associations of which one was robustly replicated at 2p16.1. Signal at 2p16.1 had lead variant rs1123573, which is in an intron of gene *BCL11A*, a zinc finger protein transcription factor involved in the regulation of the developmental switch from foetal to adult haemoglobin^64^, is essential for lymphocyte development^60^ and have previously been linked with a wide range of diseases^65^. The pleiotropic association with severe COVID-19 and IPF suggests that this gene could potentially contribute to both diseases through dysregulation of adaptive immune responses mediated by lymphocytes.

### Study limitations

Our study has several limitations:

Firstly, the GWAS summaries sourced from the GBMI meta-analysis exhibit large variations in sample size across the genome, significant heterogeneity in effect sizes due to inclusion of data from both clinical and Biobank sources^5^, and contain multi-ancestry cohorts, particularly of East Asian ancestry (Supplementary Table 7). This heterogeneity can generate local inconsistencies between summary statistics, and linkage disequilibrium computed using a European LD-reference panel may have limited our ability to identify independent signals and perform fine-mapping. For these reasons, we strictly required non-overlap of LD-clumps as well as cytogenetic loci (which encompass large genomic regions) between the current and previous studies to declare novel signals.

Secondly, to map probable effector genes for variant associations we mapped consequences for lead variants of each association with VEP and selected the gene affected or nearest gene if consequence was not genic. Our selected effector gene might not correspond to the true effector gene. Analysis on worst consequence gene in credible set and TWAS analysis integrating QTL data has highlighted additional genes that could be considered as potential effector genes to follow-up.

Thirdly, we selected *MCL1* as effector gene from the 1q21.2 novel association to follow up with *in vivo* analysis due to its intriguing connection to apoptosis and our specific hypothesis in respect to the IPF mechanism involving senescent cells. Other nearby genes in the genomic region could be potential valid targets underlying the 1q21.2 association.

## Materials and Methods

### Ethics statement

100,000 Genomes Project: the 100,000 Genomes Project was approved by the East of England-Cambridge Central Research Ethics Committee (REF 20/EE/0035). Only participants of the 100,000 Genomes Project for whom WGS data were available and who consented for their data to be used for research purposes were included in the analyses.

### IPF Cohort

The IPF cohort was constructed from participants of the 100kGP and comprised of the cases that were specifically recruited into the 100kGP for familial pulmonary fibrosis as well as ‘incidental’ cases of idiopathic pulmonary fibrosis (IPF) that were identified based on diagnostic ICD-10 codes in linked Electronic Health Records (EHRs). The incidental cases are expected to represent predominantly sporadic forms of pulmonary fibrosis but may also include a small proportion of additional familial cases.

### Sample quality control

Participants were required to have European ancestry (probability ≥ 0.8), concordance between reported phenotypic sex and WGS inferred sex, assigned either XX or XY from WGS sex chromosome ploidy checks, passed internal WGS sample QC (from BAM and VCF-level metrics) based on Blood sample, EDTA extraction, Illumina TruSeq DNA PCR-Free library preparation. Related individuals were included and for individuals with a monozygotic twin-pair (identified by KING kinship coefficient ≥ 0.354), one of the pair was excluded (cases prioritised).

### GWAS analysis

#### 100kGP GWAS site quality control

Small variants (SNPs and indels) were selected with the following site-level quality control filters: median read depth (DP) >=10, median genotype quality (GQ)>=15, ABratio for het calls >=25%, minor allele frequency (maf) > 0.5%, minor allele counts (MAC) > 20, site-wise missingness < 2%, differential missingness between cases and controls (Fisher Exact Test *P*-value < 10^−5^), Hardy Weinberg Equilibrium test for unrelated controls (HWE P-value < 10^−6^).

#### 100kGP IPF GWAS

To perform GWAS analysis in the 100kGP cohort, we used SAIGE^66^. SAIGE performs mixed model linear regression using a two-step approach where random effects for the genetic relationship matrix are estimated in step 1 (null model fit) and step 2 calculates association statistics for each variant while controlling for effects estimated in step 1. We used a set of 63,523 common (maf>5%) high-quality independent SNPs to construct the GRM and fit the null model for SAIGE part 1 (the “HQ common SNPs” from Kousathanas et al. (2022)^14^). We included sex, age, age^2^, age*sex, and the first 20 principal components calculated in Europeans as covariates in the null model fit. We then computed SAIGE summary statistics for all variants passing GWAS site QC.

#### Meta-analysis and quality control

We contacted the Global Biobank Meta-analysis Initiative (GBMI) consortium and gained access to the summary GWAS results from their latest meta-analysis^5^. This meta-analysis aggregated results from 13 Biobanks as well as another large IPF dataset described by Allen et al., 2020^3^. GBMI in-house site quality control had removed variants from each biobank with imputation quality score < 0.3 or allele frequencies differing from gnomAD. A total of 32,046,319 variants were present in the provided GBMI IPF summary statistics file, of which 10,781,762 were also present in the Allen et al. (2020)^3^ results (header column “with_Allen”). To meta-analyse with the 100kGP IPF GWAS summaries, we included variants present in a minimum of 2 GBMI biobanks (max 13) and used inverse variance weighted fixed-effect meta-analysis as implemented in METAL v.2011-03-25. The meta-analysis resulted in 10,644,973 variants that were present in both GBMI meta-analysis and the 100kGP IPF GWAS. For downstream analyses, we further removed the following variants: (i) variants with strand flips in the GBMI dataset (i.e, strand_flip=”yes”); (ii) variants with calculated allele frequency > 1 in either the GBMI or 100kGP IPF GWAS (which can arise from erroneous calculations of the number of alleles for chrX); (iii) variants with differences in absolute allele frequency between GBMI and 100kGP IPF GWAS > 0.1. These filtering steps resulted in a set of 10,287,302 variants.

#### Creation of 100kGP LD reference panel

We used an LD reference panel based on the 100kGP cohort by creating an EUR-only cohort of 44,590 unrelated individuals (LD-100kGP-EUR) and for a bi-allelic variant site subset of 9,044,108 variants. We used only Europeans for creating the LD reference panel as this ancestry represented 85% of the cases and 80% of the controls contributing to the IPF meta-analysis dataset.

#### Variant LD-clumping, conditional analysis and fine-mapping

We performed LD-clumping, conditional analysis and fine-mapping procedures using the IPF meta-analysis GWAS summary statistics and the LD-100kGP-EUR as LD-reference panel. LD-clumping was used to collect variants in LD with GWAS signals with plink1.9^67^ with *P_1_* set to 5×10^−8^, clump distance 1500kb, *P_2_*=0.01, *r^2^*=0.1.

To discover conditionally independent GWAS signals we ran a step-wise conditional analysis using GCTA 1.9.3^68^ and *–cojo-slc* function^69^. We used a *P*-value threshold of 5×10^−8^, a distance of 10,000kb and a collinear threshold of 0.1.

To narrow down the list of likely causal variants surrounding independent signals, we performed fine-mapping using R-package SusieR v0.11.42. We split the genome into 3Mbp non-overlapping windows and for each window containing at least one GW-significant variant (at *P*-value<5×10^−8^), we assigned the variant with the lowest P-value as the focal variant of the window and analysed the GWAS summaries and LD of variants within 1.5 Mbp on either side of each focal variant. This approach minimised the number of search windows for clustered signals while simultaneously centering on each signal.

#### Replication

We contacted the PFgenetics consortium (https://github.com/genomicsITER/PFgenetics) to gain access to the IPF 5-way meta-analysis summaries and 3-way meta-analysis summaries. The 3-way meta-analysis summaries had been previously included in the GBMI meta-analysis. We obtained independent IPF summary data through mathematical subtraction of the 3-way meta-analysis summaries from the 5-way meta-analysis summaries, using MetaSubtract R-package^70^. Replication of the novel signals of the present study was determined by requiring a Bonferroni-corrected P-value in the replication 2-way summaries and that the effect size direction matched.

#### Variant annotation

We used Variant Effect Predictor (VEP) v.105 for mapping variant effects. For each variant, we selected the worst predicted consequence across transcripts as the probable effector gene unless the variant was mapped as regulatory/intergenic in which case we assigned the nearest gene.

### Aggregate variant burden testing (AVT)

#### AVT site quality control

We used 100kGP aggregated data of bi-allelic variants that were processed by setting genotypes with depth of coverage (DP) < 10, genotype quality (GQ) < 20 and heterozygote genotypes failing an ABratio binomial test with *P* value < 10^−3^ to missing with bcftools *setGT* module. This was followed by a site quality control procedure that removed sites with >2% missingness and performed a Fisher’s exact test of missingness between cases and controls and filtered-out variants with *P*-value<10^−5^.

#### AVT model

Aggregate variant burden testing (AVT) was performed using SAIGE-GENE+ (version 0.44.6.5)^71^. Sex, age, age^2^, age*sex, and the first 20 principal components calculated in Europeans were included as covariates in the SAIGE-GENE+ null model fit.

#### AVT masks

Small variants (SNPs and indels) were annotated using Ensembl VEP^72^ and a suite of additional functional and allele frequency datasets (including CADD, LOFTEE, and gnomAD). Variants annotated against all protein-coding transcripts from Ensembl VEP 105 (including canonical and non-canonical transcripts) were aggregated by gene and grouped into four variant masks shown in Supplementary Table 11. These masks were selected to align with existing AVT studies in IPF ^8,19,73^.

### Post-GWAS analyses

#### Transcriptome-wide association study (TWAS)

We conducted a transcriptome-wide association study (TWAS) for lung tissue using the MetaXcan framework^20^ along with the GTEx v.8 (eQTL) MASHR-M model (downloaded from http://predictdb.org/) using the S-PrediXcan function. We identified significant genes using the Bonferroni correction.

#### Colocalisation for TWAS

We tested for colocalisation between IPF and cis-eQTL summary statistics for genes with significant TWAS results. For each significant TWAS gene, we selected eQTLs located 1.5 Mbp on either side of the transcription start site (TSS) of the gene (as defined in Ensembl v104). We used Bayesian colocalisation analysis utilising the *coloc_abf* function implemented in R-package coloc v5.1.0^23^. We declared that a locus had significant evidence for colocalisation when both of two criteria were met: (1) At *p_12_*=5×10^−5^, the probability of colocalisation with a shared causal variant (*PP_H4_*) was > 0.5, and (2) at *p_12_*=10^−5^, the probability of distinct causal signals (*PP_H3_*) was not the main hypothesis (*PP_H3_* < 0.5). For each locus we determined the lead colocalisation variant as the one with the highest *PP_H4_*.

#### Colocalisation for IPF and severe-COVID-19

We tested for colocalization of each genome-wide significant IPF signal with severe COVID-19 by selecting variants located 1.5Mbp on either side of the lead variant and performing colocalisation analysis using coloc v5.1.0 as for TWAS described above.

#### Genetic correlation

To compute genetic correlations between IPF and COVID-19 we used LDSC (https://github.com/bulik/ldsc)^74^. For this analysis, we used pre-computed European ancestry LD-score weights for GRCh38 coordinates. We used the files with prefix “weights.hm3_noMHC” that are split by chromosome. Running LDSC was performed in two steps: (1) Cleaning the summary statistics of each analysed trait for flip strands and taking an intersection with the LD-scores variants with script *munge_sumstats.py*, (2) Running genetic correlation analysis on pairs of traits with script *ldsc.py*.

#### Multi-trait meta-analysis (MTAG)

To perform multi-trait meta-analysis between IPF and COVID-19 we used MTAG (https://github.com/JonJala/mtag)^24^. For this analysis, we used European ancestry pre-computed LD-score weights for GRCh38 coordinates. We used the files with prefix “weights.hm3_noMHC” that are split by chromosome. We performed variant clumping to collect variants in LD with GWAS signals and to help identify novel signals compared to the single trait meta-analysis, a process that resulted in 71 MTAG-derived significant LD-clumps for IPF. Due to the genetic correlation between IPF and severe COVID-19 being only moderate and the MTAG assumption for a constant genetic correlation across the genome, we expected that many signals in either trait will be entirely driven by strong signals of the paired rather than the focal trait. Therefore, to reduce the number of false positives when declaring significance, we required that the *P*-value of the focal SNP in each MTAG IPF LD-clump was also significant in the original single trait meta-analysis at a Bonferroni corrected threshold for the number of clumps in the MTAG results (0.05 / 71 = 7.04×10^−4^), and that the effect sizes were consistent between the original and the MTAG meta-analyses. 13 LD-clumps passed these criteria and we identified novel signals as those occurring in non-overlapping LD-clumps as well as different loci compared to the base IPF meta-analysis.

## Competing Interests

Genomics England is a limited company that is wholly owned by the UK Department of Health and Social Care, established in 2013 to run the 100,000 Genomes Project and introduce whole genome sequencing and advanced analytics into the NHS to evolve genomic healthcare. All Genomics England affiliated authors are, or were, salaried by Genomics England during this programme. Authors affiliated with Boehringer Ingelheim Pharma GmbH & Co. KG are all current employees of Boehringer Ingelheim. All other authors declare that they have no competing interests relating to this work.

## Data Availability

All data produced in the present study are available upon reasonable request to the authors

## Notes

### Funding Statement

This study did not receive any funding

## References

1. Fujimoto, H., Kobayashi, T. & Azuma, A. Idiopathic Pulmonary Fibrosis: Treatment and Prognosis. Clin Med Insights Circ Respir Pulm Med 9, 179 (2015).

2. Seibold, M. A. et al. A Common MUC5B Promoter Polymorphism and Pulmonary Fibrosis. New England Journal of Medicine 364, 1503–1512 (2011).

3. Allen, R. J. et al. Genome-Wide Association Study of Susceptibility to Idiopathic Pulmonary Fibrosis. Am J Respir Crit Care Med 201, 564–574 (2020).

4. Allen, R. J. et al. Genome-wide association study across five cohorts identifies five novel loci associated with idiopathic pulmonary fibrosis. Thorax 77, 829 (2022).

5. Partanen, J. J. et al. Leveraging global multi-ancestry meta-analysis in the study of idiopathic pulmonary fibrosis genetics. Cell Genomics 2, 100181 (2022).

6. Petrovski, S. et al. An Exome Sequencing Study to Assess the Role of Rare Genetic Variation in Pulmonary Fibrosis. Am J Respir Crit Care Med 196, 82–93 (2017).

7. Peljto, A. L. et al. Idiopathic Pulmonary Fibrosis Is Associated with Common Genetic Variants and Limited Rare Variants. Am J Respir Crit Care Med 207, (2023).

8. Dhindsa, R. S., et al. Identification of a missense variant in SPDL1 associated with idiopathic pulmonary fibrosis. Commun Biol 4, (2021).

9. Zhang, D. et al. Rare and Common Variants in KIF15 Contribute to Genetic Risk of Idiopathic Pulmonary Fibrosis. Am J Respir Crit Care Med 206, 56–69 (2022).

10. Molyneaux, P. L. & Maher, T. M. The role of infection in the pathogenesis of idiopathic pulmonary fibrosis. European Respiratory Review 22, 376–381 (2013).

11. Fadista, J. et al. Shared genetic etiology between idiopathic pulmonary fibrosis and COVID-19 severity. EBioMedicine 65, (2021).

12. Allen, R. J. et al. Genetic overlap between idiopathic pulmonary fibrosis and COVID−19. European Respiratory Journal 60, (2022).

13. Pairo-Castineira, E. et al. GWAS and meta-analysis identifies 49 genetic variants underlying critical COVID-19. Nature 2023 617:7962 617, 764–768 (2023).

14. Kousathanas, A. et al. Whole-genome sequencing reveals host factors underlying critical COVID-19. Nature 2022 607:7917 607, 97–103 (2022).

15. Niemi, M. E. K. et al. Mapping the human genetic architecture of COVID-19. Nature 2021 600:7889 600, 472–477 (2021).

16. Seibold, M. A. et al. A common MUC5B promoter polymorphism and pulmonary fibrosis. N Engl J Med 364, 1503–1512 (2011).

17. Yang, J., Lee, S. H., Goddard, M. E. & Visscher, P. M. GCTA: a tool for genome-wide complex trait analysis. Am J Hum Genet 88, 76–82 (2011).

18. Wang, G., Sarkar, A., Carbonetto, P. & Stephens, M. A simple new approach to variable selection in regression, with application to genetic fine mapping. J R Stat Soc Series B Stat Methodol 82, 1273–1300 (2020).

19. Petrovski, S. et al. An Exome Sequencing Study to Assess the Role of Rare Genetic Variation in Pulmonary Fibrosis. Am J Respir Crit Care Med 196, 82–93 (2017).

20. Barbeira, A. N. et al. Exploring the phenotypic consequences of tissue specific gene expression variation inferred from GWAS summary statistics. Nature Communications 2018 9:1 9, 1–20 (2018).

21. Chen, M. et al. Integrative analyses for the identification of idiopathic pulmonary fibrosis-associated genes and shared loci with other diseases. Thorax 78, 792–798 (2023).

22. Gong, W., Guo, P., Liu, L., Guan, Q. & Yuan, Z. Integrative Analysis of Transcriptome-Wide Association Study and mRNA Expression Profiles Identifies Candidate Genes Associated With Idiopathic Pulmonary Fibrosis. Front Genet 11, (2020).

23. Giambartolomei, C. et al. Bayesian Test for Colocalisation between Pairs of Genetic Association Studies Using Summary Statistics. PLoS Genet 10, e1004383 (2014).

24. Turley, P. et al. Multi-trait analysis of genome-wide association summary statistics using MTAG. Nat Genet 50, 229–237 (2018).

25. Initiative, T. C.-19 H. G. & Ganna, A. A second update on mapping the human genetic architecture of COVID-19. medRxiv 2022.12.24.22283874 (2023) doi:10.1101/2022.12.24.22283874.

26. Habermann, A. C. et al. Single-cell RNA sequencing reveals profibrotic roles of distinct epithelial and mesenchymal lineages in pulmonary fibrosis. Sci Adv 6, (2020).

27. Campisi, J. & D’Adda Di Fagagna, F. Cellular senescence: when bad things happen to good cells. Nat Rev Mol Cell Biol 8, 729–740 (2007).

28. Cooley, J. C., et al. Inhibition of antiapoptotic BCL-2 proteins with ABT-263 induces fibroblast apoptosis, reversing persistent pulmonary fibrosis. JCI Insight 8, (2023).

29. Schafer, M. J. et al. Cellular senescence mediates fibrotic pulmonary disease. Nat Commun 8, (2017).

30. Merkt, W., Bueno, M., Mora, A. L. & Lagares, D. Senotherapeutics: Targeting senescence in idiopathic pulmonary fibrosis. Seminars in Cell and Developmental Biology vol. 101 Preprint at 10.1016/j.semcdb.2019.12.008 (2020).

31. Delbridge, A. R. D., Grabow, S., Strasser, A. & Vaux, D. L. Thirty years of BCL-2: translating cell death discoveries into novel cancer therapies. Nat Rev Cancer 16, 99– 109 (2016).

32. Meiners, S. & Lehmann, M. Senescent Cells in IPF: Locked in Repair? Front Med (Lausanne) 7, (2020).

33. Han, S., Lu, Q. & Liu, X. Advances in cellular senescence in idiopathic pulmonary fibrosis (Review). Exp Ther Med 25, (2023).

34. Cooley, J. C., et al. Inhibition of antiapoptotic BCL-2 proteins with ABT-263 induces fibroblast apoptosis, reversing persistent pulmonary fibrosis. JCI Insight 8, (2023).

35. Merkt, W., Bueno, M., Mora, A. L. & Lagares, D. Senotherapeutics: Targeting senescence in idiopathic pulmonary fibrosis. Seminars in Cell and Developmental Biology vol. 101 Preprint at 10.1016/j.semcdb.2019.12.008 (2020).

36. Schafer, M. J. et al. Cellular senescence mediates fibrotic pulmonary disease. Nat Commun 8, (2017).

37. Mohamad Anuar, N. N., Nor Hisam, N. S., Liew, S. L. & Ugusman, A. Clinical Review: Navitoclax as a Pro-Apoptotic and Anti-Fibrotic Agent. Frontiers in Pharmacology vol. 11 Preprint at 10.3389/fphar.2020.564108 (2020).

38. Hafezi, S. & Rahmani, M. Targeting bcl-2 in cancer: Advances, challenges, and perspectives. Cancers vol. 13 Preprint at 10.3390/cancers13061292 (2021).

39. Negi, A. & Murphy, P. V. Development of Mcl-1 inhibitors for cancer therapy. European Journal of Medicinal Chemistry vol. 210 Preprint at 10.1016/j.ejmech.2020.113038 (2021).

40. van Geffen, C. et al. Regulatory Immune Cells in Idiopathic Pulmonary Fibrosis: Friends or Foes? Front Immunol 12, (2021).

41. Venosa, A. Senescence in Pulmonary Fibrosis: Between Aging and Exposure. Front Med (Lausanne) 7, 606462 (2020).

42. Tanigawa, Y. et al. Rare protein-altering variants in ANGPTL7 lower intraocular pressure and protect against glaucoma. PLoS Genet 16, (2020).

43. Camenisch, G. et al. ANGPTL3 stimulates endothelial cell adhesion and migration via integrin alpha vbeta 3 and induces blood vessel formation in vivo. J Biol Chem 277, 17281–17290 (2002).

44. Zheng, J. et al. Inhibitory receptors bind ANGPTLs and support blood stem cells and leukaemia development. Nature 485, 656–660 (2012).

45. Ben-Zvi, D. et al. Angptl4 links α-cell proliferation following glucagon receptor inhibition with adipose tissue triglyceride metabolism. Proc Natl Acad Sci U S A 112, 15498–15503 (2015).

46. Praveen, K. et al. ANGPTL7, a therapeutic target for increased intraocular pressure and glaucoma. Commun Biol 5, 1051 (2022).

47. Toyono, T. et al. Angiopoietin-like 7 is an anti-angiogenic protein required to prevent vascularization of the cornea. PLoS One 10, (2015).

48. Parri, M. et al. Angiopoietin-like 7, a novel pro-angiogenetic factor over-expressed in cancer. Angiogenesis 17, 881–896 (2014).

49. Neumark, N., Cosme, C., Rose, K. A. & Kaminski, N. The Idiopathic Pulmonary Fibrosis Cell Atlas. Am J Physiol Lung Cell Mol Physiol 319, L887–L893 (2020).

50. Liew, C. W. et al. Ablation of TRIP-Br2, a novel regulator of fat lipolysis, thermogenesis and oxidative metabolism, prevents diet-induced obesity and insulin resistance. Nat Med 19, 217 (2013).

51. Cheong, J. K. et al. TRIP-Br2 promotes oncogenesis in nude mice and is frequently overexpressed in multiple human tumors. J Transl Med 7, (2009).

52. Chen, Y. et al. REV1 promotes lung tumorigenesis by activating the Rad18/SERTAD2 axis. Cell Death Dis 13, (2022).

53. Melià, M. J. et al. Limb-girdle muscular dystrophy 1F is caused by a microdeletion in the transportin 3 gene. Brain 136, 1508 (2013).

54. Costa, R. et al. Transportin 3 (TNPO3) and related proteins in limb girdle muscular dystrophy D2 muscle biopsies: A morphological study and pathogenetic hypothesis. Neuromuscul Disord 30, 685–692 (2020).

55. Zochling, J. et al. An Immunochip-based interrogation of scleroderma susceptibility variants identifies a novel association at DNASE1L3. Arthritis Res Ther 16, (2014).

56. López-Isac, E. et al. GWAS for systemic sclerosis identifies multiple risk loci and highlights fibrotic and vasculopathy pathways. Nat Commun 10, (2019).

57. Pan, P. et al. SARS-CoV-2 N protein enhances the anti-apoptotic activity of MCL-1 to promote viral replication. Signal Transduct Target Ther 8, (2023).

58. Lee, S. et al. Virus-induced senescence is a driver and therapeutic target in COVID-19. Nature 2021 599:7884 599, 283–289 (2021).

59. Mathai, S. K. et al. Desmoplakin Variants Are Associated with Idiopathic Pulmonary Fibrosis. Am J Respir Crit Care Med 193, 1151–1160 (2016).

60. Yu, Y. et al. Bcl11a is essential for lymphoid development and negatively regulates p53. J Exp Med 209, 2467 (2012).

61. Burzenski, L. M. et al. Inactive rhomboid proteins RHBDF1 and RHBDF2 (iRhoms): a decade of research in murine models. Mammalian Genome 32, 415 (2021).

62. Razzaq, A. et al. Targeting epidermal growth factor receptor signalling pathway: A promising therapeutic option for COVID-19. Rev Med Virol 34, e2500 (2024).

63. Turley, P. et al. Multi-trait analysis of genome-wide association summary statistics using MTAG. Nat Genet 50, 229–237 (2018).

64. Liu, N. et al. Direct Promoter Repression by BCL11A Controls the Fetal to Adult Hemoglobin Switch. Cell 173, 430–442.e17 (2018).

65. Yin, J., Xie, X., Ye, Y., Wang, L. & Che, F. BCL11A: a potential diagnostic biomarker and therapeutic target in human diseases. Biosci Rep 39, (2019).

66. Zhou, W. et al. Efficiently controlling for case-control imbalance and sample relatedness in large-scale genetic association studies. Nature Genetics 2018 50:9 50, 1335–1341 (2018).

67. Purcell, S. et al. PLINK: A Tool Set for Whole-Genome Association and Population-Based Linkage Analyses. Am J Hum Genet 81, 559 (2007).

68. Yang, J., Lee, S. H., Goddard, M. E. & Visscher, P. M. GCTA: a tool for genome-wide complex trait analysis. Am J Hum Genet 88, 76–82 (2011).

69. Yang, J. et al. Conditional and joint multiple-SNP analysis of GWAS summary statistics identifies additional variants influencing complex traits. Nat Genet 44, 369 (2012).

70. Nolte, I. M. Metasubtract: an R-package to analytically produce leave-one-out meta-analysis GWAS summary statistics. Bioinformatics 36, 4521–4522 (2020).

71. Zhou, W. et al. SAIGE-GENE+ improves the efficiency and accuracy of set-based rare variant association tests. Nature Genetics 2022 54:10 54, 1466–1469 (2022).

72. McLaren, W. et al. The Ensembl Variant Effect Predictor. Genome Biol 17, 1–14 (2016).

73. Zhao, H. et al. Proteome-wide Mendelian randomization in global biobank meta-analysis reveals multi-ancestry drug targets for common diseases. Cell Genomics 2, 100195 (2022).

74. Bulik-Sullivan, B. et al. LD Score Regression Distinguishes Confounding from Polygenicity in Genome-Wide Association Studies. Nat Genet 47, 291 (2015).

