## Supplementary Material for "Common and rare variant analyses reveal novel genetic factors underlying Idiopathic Pulmonary Fibrosis and shared aetiology with COVID-19"

### 100,000 Genomes Project (100kGP) cohorts

#### 100kGP cohorts definition

*Supplementary Table 1. Case control definition for the 100kGP FPF and IPF cohorts*

<table>
<tr><th>Cohort</th><th>Case Definition</th><th>Control Definition (Exclusion Criteria)</th></tr>
<tr><td>FPF</td><td>Recruited Rare Disease Term: Familial Pulmonary Fibrosis.</td><td rowspan="2">Recruited Rare Disease Group: Respiratory Disorders OR Positive for HPO term: HP:0002086 (Abnormality of the respiratory system) OR Positive for any descendent HPO terms of HP:0002086 (n=319) OR Any ICD-10 Diagnosis from EHRs derived from Phecodes 500-509.99* (Pneumoconiosis and other lung diseases) OR Positive for Recruited Cancer: Lung</td></tr>
<tr><td>IPF</td><td>Recruited Rare Disease term: Familial Pulmonary Fibrosis OR Positive for HPO term: HP:0002206 (pulmonary fibrosis) OR Any ICD-10 Diagnosis from EHRs of: J84.1, (Other interstitial pulmonary diseases with fibrosis) J84.8 (Other specified interstitial pulmonary diseases), J84.9 (Interstitial pulmonary disease, unspecified).</td></tr>
</table>

** Exclusion Phecodes 500-509.99 include: Lung disease due to external agents, Extrinsic allergic alveolitis, Pneumoconiosis, Pneumonitis due to inhalation of food or vomitus, Post-inflammatory pulmonary fibrosis, Pulmonary congestion and hypostasis, Other alveolar and parietoalveolar pneumopathy, Other pulmonary inflammation or oedema, Empyema and pneumothorax, Pleurisy; pleural effusion, Pulmonary collapse; interstitial and compensatory emphysema, Respiratory failure, Respiratory insufficiency, Pulmonary insufficiency or respiratory failure following trauma and surgery, Respiratory arrest, Dependence on respirator [Ventilator] or supplemental oxygen. Available from https://phewascatalog.org/phecodes_icd10.*

*Supplementary Table 2. 100kGP Recruitment Criteria, panel of HPO terms assessed, and genes tested for Familial Pulmonary Fibrosis (FPF) as part of the Genomics England interpretation pipeline.*

| Eligibility Criteria | Description |
|---|---|
| **FPF inclusion criteria** | • Clinical syndrome consistent with interstitial lung disease: Breathlessness on exertion or cough, bilateral crepitations on examination, AND<br>• A High Resolution CT scan with evidence of interstitial lung disease, AND<br>• A first degree relative with an interstitial lung disease |
| **FPF exclusion criteria** | • A respiratory disease other than an interstitial lung disease<br>• Any cystic lung disease |
| **Prior genetic testing guidance** | Results should have been reviewed for all genetic tests undertaken, including disease-relevant genes in exome sequencing data. The patient is not eligible if they have a molecular diagnosis for their condition.<br>Genetic testing should continue according to routine local practice for this phenotype regardless of recruitment to the project; results of these tests must be submitted via the Genetic investigations section of the data capture tool to allow comparison of WGS with current standard testing.<br>PLEASE NOTE: The sensitivity of WGS compared to current diagnostic genetic testing has not yet been established. It is therefore important that tests which are clinically indicated under local standard practice continue to be carried out. |
| **FPF prior genetic testing genes** | Testing of the following genes should be carried out PRIOR TO RECRUITMENT where this is in line with current local practice:<br>• SFTPB, SFTPC in childhood onset cases |
| **FPF Panel of HPO Terms Assessed** | Interstitial pulmonary abnormality (HP:0006530), Pulmonary fibrosis (HP:0002206), Acute respiratory tract infection (HP:0011948), Asthma (HP:0002099), Tachypnea (HP:0002789), Dyspnea (HP:0002094), Cough (HP:0012735), Hemoptysis (HP:0002105), Aspiration (HP:0002835), Hypertension (HP:0000822), Angina pectoris (HP:0001681), Coronary artery disease (HP:0001677), Stroke (HP:0001297), Diabetes mellitus (HP:0000819), Deep venous thrombosis (HP:0002625), Pulmonary embolism (HP:0002204), Neoplasm (HP:0002664), Renal insufficiency (HP:0000083), Thromboembolism (HP:0001907), Rheumatoid arthritis (HP:0001370), Osteoarthritis (HP:0002758), Depression (HP:0000716), Anxiety (HP:0000739), Gastroesophageal reflux (HP:0002020) |
| **FPF Green Genes PanelApp (n=26) version 1.30** | *ABCA3, ACD, AP3B1, ASAH1, CSF2RA, CSF2RB, DKC1, FAM111B, FARSA, FARSB, GBA, HPS1, HPS4, ITGA3, NKX2-1, PARN, RTEL1, SFTPA2, SFTPB, SFTPC, SLC34A2, SLC7A7, SMPD1, TERC, TERT, TINF2* |

#### 100kGP FPF solved cases

All 147 FPF participants were processed through the Genomics England Rare Disease Interpretation Pipeline - which aims to assess plausibly pathogenic variants and enable genetic diagnosis for participants.

The Interpretation Pipeline annotates variants based on their segregation in the family, frequency in control populations, effect on protein coding genes, mode of inheritance, and whether they are in a gene in the virtual gene panel(s) applied to the family. TIER 1 variants represent protein truncating or *de novo* variants in 'green' genes (genes with high confidence of disease association - see PanelApp [https://panelapp.genomicsengland.co.uk/]). TIER 2 variants represent protein-altering variants (e.g. missense) in green genes.

Only six FPF participants (4%) were assigned a TIER 1 variant (Supplementary Table 3) and were marked as 'Case Solved' by the Genomic Medicine Centre clinician (Supplementary Table 4). These six participants were deemed to have pathogenic variants in one of the following four genes: *PARN*, *RTEL1*, *TERT*, *COPA*. Note that the case solved rate across all rare diseases in the 100kGP is approximately 20%; highlighting that the majority of FPF cases do not have high impact variants in known FPF disease genes.

*Supplementary Table 3. Number of participants with Tiered variants in the FPF cohort*

| TIER | FPF Participants | Percent of all FPF Participants |
|---|---|---|
| **TIER 1** | 6 | 4% |
| **TIER 2** | 28 | 19% |

*Supplementary Table 4. Number of Case Solved participants in the FPF cohort*

| FPF Cases Solved | FPF Cases Unsolved | Percent FPF Solve Rate |
|---|---|---|
| 6 | 141 | 4% |

#### 100kGP cohort demographics

The following tables and figures show a breakdown of the FPF and IPF cohorts by: age (Supplementary Table 5 and Supplementary Figure 1), sex (Supplementary Table 6) and principal components 1-4 projections (Supplementary Figure 2). Age refers to age at time of recruitment into the 100kGP (approximately the age at which WGS was performed). Sex refers to the reported phenotypic sex stated at recruitment. Cases are labelled as '1' and controls as '0'.

*Supplementary Table 5. Age summary at time of recruitment into 100kGP*

| Cohort | Case / Control | Participants | Mean (SD) |
|---|---|---|---|
| **FPF** | 0 | 52,084 | 42.1 (21.1) |
| **FPF** | 1 | 147 | 66.3 (14.0) |
| **IPF** | 0 | 52,083 | 42.1 (21.1) |
| **IPF** | 1 | 586 | 64.0 (17.6) |

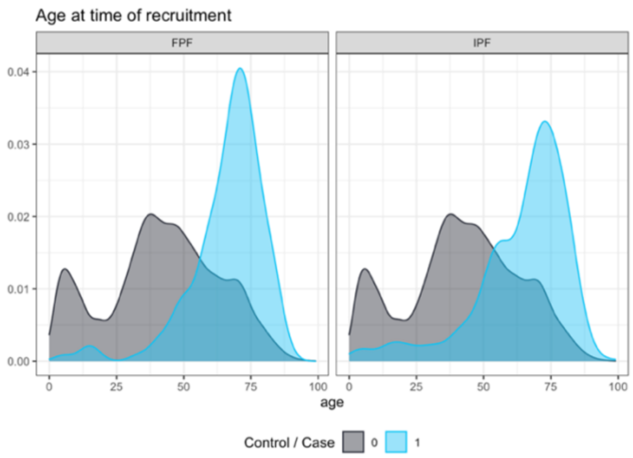

*Supplementary Figure 1. Density plot of age at time of recruitment into 100kGP of IPF and FPF cohorts*

*Supplementary Table 6. Reported phenotypic sex*

| Cohort | Case / Control | Participants | Male | Female | Male (%) | Female (%) |
|---|---|---|---|---|---|---|
| **FPF** | 0 | 52,084 | 28,373 | 23,711 | 54.5 | 45.5 |
| **FPF** | 1 | 147 | 79 | 68 | 53.7 | 46.3 |
| **IPF** | 0 | 52,083 | 28,372 | 23,711 | 54.5 | 45.5 |
| **IPF** | 1 | 586 | 293 | 293 | 50 | 50 |

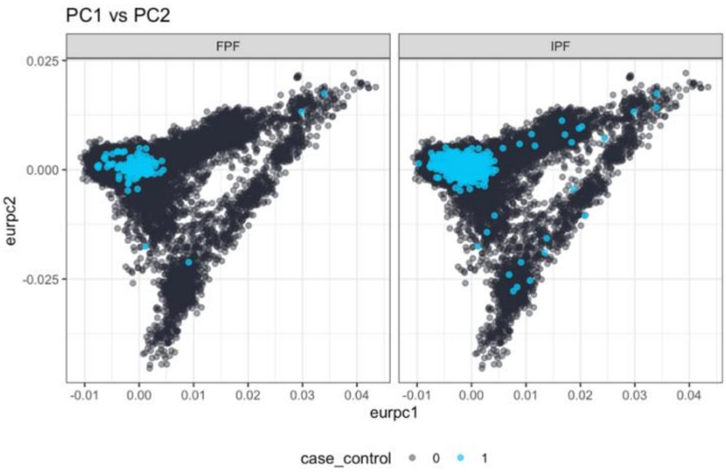

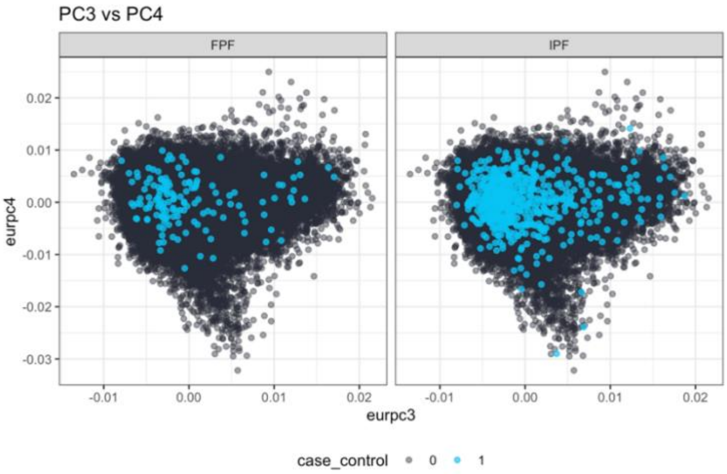

*Supplementary Figure 2. PCs 1-4 of IPF & FPF control cohorts coloured by case / control status.*

### GWAS analysis

#### 100kGP GWAS

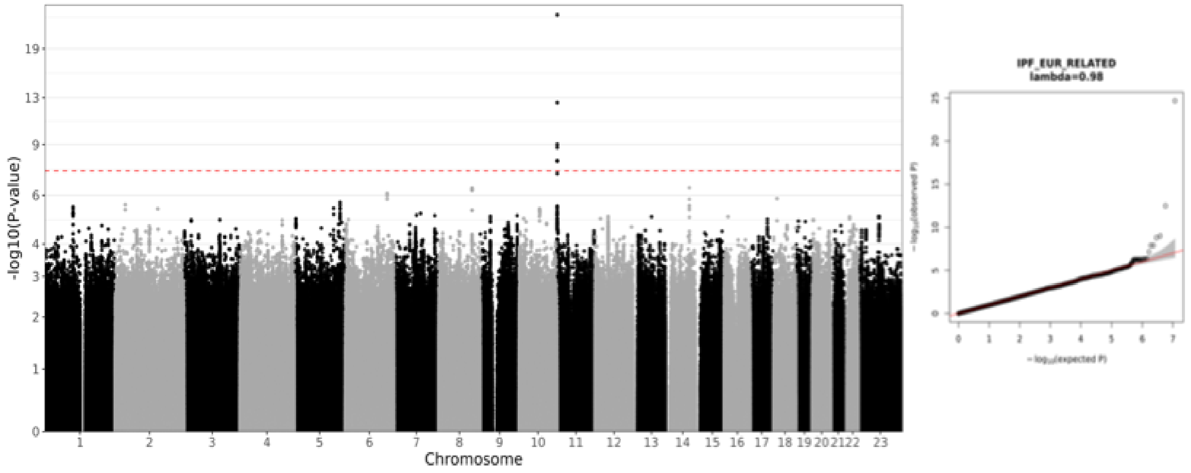

*Supplementary Figure 3. Manhattan and Q-Q plots for the 100,000 Genomes Project (100kGP) GWAS discovery analysis on 586 IPF cases and 52,083 controls of European ancestry.*

#### Meta-analysis GWAS

*Supplementary Table 7. Case and control counts and ancestry breakdown of meta-analysed IPF cohorts.*

| Cohort | Ancestry | IPF Cases | Controls | Total |
|---|---|---|---|---|
| **BioVU, CCPM, ESTBB, HUNT, MGB, MGI, UCLA, UKBB** | NFE | 5,229 | 750,630 | 755,859 |
| **FinnGen** | FIN | 1,514 | 306,063 | 307,577 |
| **BBJ, CKB, UCLA** | EAS | 1,210 | 254,409 | 255,619 |
| **BioMe, UCLA** | AMR | 319 | 14,452 | 14,771 |
| **BioMe, UCLA** | AFR | 169 | 8,368 | 8,537 |
| **GNH** | SAS | 51 | 21,897 | 21,948 |
| **GBMI [Includes all above]** | multiple | 8,492 | 1,355,819 | 1,364,311 |
| **Allen et al. (2020)** | EUR | 2,668 | 8,591 | 11,259 |
| **GBMI + Allen et al. (2020) [Partanen et al. (2022]** | multiple | 11,160 | 1,364,410 | 1,375,570 |
| **100kGP (This study)** | EUR | 586 | 52,083 | 52,669 |
| **Partanen et al. (2022) + 100kGP (This study)** | multiple | 11,746 | 1,416,493 | 1,428,239 |

NFE=Non-Finnish Europeans, FIN=Finnish, EAS=East Asian, AMR=American, AFR=African, SAS=South Asian, EUR=Europeans

*Supplementary Table 8. Independent signals of IPF meta-analysis reaching genome-wide significance as identified by GCTA-COJO from the analysis of 9,044,108 bi-allelic variants. Loci are displayed as cytogenetic band locations of the variants, allowing each locus to harbour multiple independent signals. For each variant, two IDs are shown: The variant 100kGP ID consists of Chromosome, position in hg38 coordinates and reference and alternate hg38 alleles (chr:pos_ref_alt), and we also give the respective rsID. AF EUR corresponds to the allele frequency of the variants in a set of N=44,590 unrelated European individuals from the 100kGP that was used also as the LD reference for the conditional analysis. Odds ratios (OR) are calculated for the alternate allele. Nvar clump corresponds to the number of variants included in the LD clump that the displayed variant belongs. VEP v.105-defined most severe consequence across transcripts and the impacted gene is shown. The nearest gene in terms of the closest distance to the Transcription Start Site (TSS) for each variant is shown.* ***Blue font*** *identifies novel locus signals of this study compared to Partanen et al. (2022)*[1].

| Locus | Variant ID chr:pos_ref_alt | rsID | AF EUR | OR (95%CI) | *P*-value | Nvar clump | VEP most severe Consequence | VEP gene | Nearest gene |
|---|---|---|---|---|---|---|---|---|---|
| **1p36.23** | chr1:9107187_C_A | rs7549256 | 0.68 | 0.91 (0.89, 0.94) | 6.98E-09 | 80 | intron | *GPR157* | *SLC2A5* |
| **1p31.1** | chr1:77998184_T_C | rs4130548 | 0.38 | 1.09 (1.06, 1.12) | 3.51E-08 | 109 | Intron | *DNAJB4* | *GIPC2* |

| | | | | | | | | | |
|---|---|---|---|---|---|---|---|---|---|
| **1q21.2** | **chr1:150579566_G_A** | **rs16837903** | **0.15** | **0.88 (0.85, 0.92)** | **9.54E-09** | **239** | **5' UTR** | ***MCL1*** | ***MCL1*** |
| **3p21.31** | chr3:44804157_T_C | rs74341405 | 0.042 | 1.27 (1.19, 1.36) | 1.40E-12 | 91 | Intron | *KIF15* | *KIF15* |
| **3q26.2** | chr3:169774313_C_T | rs10936599 | 0.24 | 1.16 (1.12, 1.19) | 6.37E-20 | 144 | Synonymous | *MYNN* | *MYNN* |
| **4q22.1** | chr4:88915668_C_T | rs2609260 | 0.82 | 0.85 (0.82, 0.88) | 3.53E-21 | 172 | Intron | *FAM13A* | *FAM13A* |
| **5p15.33** | chr5:1282299_G_A | rs7725218 | 0.34 | 0.82 (0.79, 0.84) | 3.39E-38 | 14 | Intron | *TERT* | *TERT* |
| **5p15.33** | chr5:1442411_G_A | rs2652514 | 0.26 | 1.11 (1.08, 1.15) | 1.58E-10 | 29 | Intron | *SLC6A3* | *SLC6A3* |
| **5q35.1** | chr5:169588475_G_A | rs116483731 | 0.0074 | 1.94 (1.71, 2.21) | 2.25E-24 | 13 | Missense | *SPDL1* | *SPDL1* |
| **6p24.3** | chr6:7562999_T_G | rs2076295 | 0.45 | 1.21 (1.18, 1.25) | 9.00E-43 | 41 | Intron | *DSP* | *DSP* |
| **6p21.33** | chr6:31380300_G_A | rs9266669 | 0.16 | 1.13 (1.09, 1.18) | 6.12E-10 | 1131 | Upstream gene | *ZDHHC20P2* | *HLA-B* |
| **7p22.3** | chr7:1937227_G_A | rs2280546 | 0.40 | 0.91 (0.89, 0.94) | 1.04E-09 | 437 | Intron | *MAD1L1* | *ELFN1* |
| **7q22.1** | chr7:100007746_G_A | rs11770546 | 0.37 | 1.16 (1.12, 1.19) | 1.06E-23 | 244 | Intron | - | *ZKSCAN1* |
| **7q32.1** | chr7:129095384_G_A | rs34288126 | 0.13 | 1.13 (1.08, 1.18) | 1.42E-08 | 106 | Non_coding transcript_exon | - | *TNPO3* |
| **8q22.1** | **chr8:97822998_T_C** | **rs6983263** | **0.48** | **0.92 (0.9, 0.95)** | **4.47E-08** | **201** | **Intron** | ***LAPTM4B*** | ***MATN2*** |
| **9q31.3** | chr9:110842172_T_C | rs77880635 | 0.04 | 1.16 (1.1, 1.23) | 3.56E-08 | 9 | Regulatory region | - | *MUSK* |
| **11p15.5** | chr11:1013992_C_T | rs148815783 | 0.0073 | 2.1 (1.81, 2.44) | 8.58E-23 | 3 | Missense | *MUC6* | *MUC6* |
| **11p15.5** | chr11:1057948_G_C | rs12574439 | 0.12 | 1.21 (1.15, 1.28) | 2.56E-13 | 36 | Downstream gene | *LINC02688* | *MUC2* |
| **11p15.5** | chr11:1078249_C_T | rs7104590 | 0.69 | 1.15 (1.11, 1.19) | 3.89E-16 | 2 | Intron | *MUC2* | *MUC2* |
| **11p15.5** | chr11:1081750_C_T | rs41359951 | 0.018 | 1.55 (1.41, 1.7) | 1.29E-20 | 2 | Synonymous | *MUC2* | *MUC2* |
| **11p15.5** | chr11:1208601_C_T | rs548688097 | 0.0070 | 2.84 (2.36, 3.41) | 4.09E-29 | 6 | Intergenic | - | *MUC5B* |
| **11p15.5** | chr11:1214934_A_G | rs12802931 | 0.20 | 1.71 (1.63, 1.78) | 3.35E-123 | 88 | Upstream_gene | - | *MUC5B* |
| **11p15.5** | chr11:1219775_T_C | rs2672794 | 0.61 | 1.2 (1.16, 1.25) | 2.87E-26 | 38 | Intron | - | *MUC5B* |
| **11p15.5** | chr11:1224068_C_T | rs56321310 | 0.19 | 1.39 (1.34, 1.44) | 2.71E-67 | 96 | Intron | *MUC5B* | *MUC5B* |
| **11p15.5** | chr11:1289602_C_T | rs34787245 | 0.012 | 2.97 (2.52, 3.5) | 2.55E-39 | 5 | Intron | *TOLLIP* | *TOLLIP* |
| **13q34** | chr13:112881427_C_T | rs12585036 | 0.21 | 0.89 (0.86, 0.92) | 2.22E-11 | 57 | Non_coding transcript_exon | *ATP11A* | *MCF2L* |
| **15q15.1** | chr15:40428343_G_T | rs59424629 | 0.52 | 1.16 (1.13, 1.19) | 4.28E-24 | 96 | Intron | *IVD* | *BAHD1* |
| **15q15.1** | chr15:40677024_C_G | rs7176221 | 0.85 | 0.87 (0.84, 0.9) | 2.00E-12 | 80 | intergenic | - | *RAD51* |
| **15q25.3** | chr15:85748038_C_T | rs3169119 | 0.25 | 1.13 (1.09, 1.16) | 2.96E-11 | 602 | 3' UTR | *AKAP13* | *KLHL25* |
| **16p13.3** | chr16:320936_C_T | rs564373006 | 0.0051 | 1.65 (1.39, 1.95) | 1.15E-08 | 1 | Intron | *AXIN1* | *AXIN1* |
| **17q21.31** | chr17:46126154_C_T | rs113120855 | 0.22 | 0.8 (0.77, 0.84) | 1.48E-25 | 868 | Intron | *KANSL1* | *KANSL1* |
| **19p13.3** | chr19:4717660_A_G | rs12610495 | 0.30 | 1.11 (1.08, 1.15) | 4.38E-11 | 33 | non_coding transcript_exon | *DPP9* | *DPP9* |
| **19p13.3** | chr19:5847989_T_C | rs2608894 | 0.76 | 0.9 (0.87, 0.94) | 1.51E-08 | 43 | Intron | *FUT3* | *FUT3* |
| **20q13.33** | chr20:63623343_A_G | rs148583745 | 0.018 | 1.42 (1.31, 1.55) | 1.17E-15 | 58 | Intron | *GMEB2* | *GMEB2* |

### Post-GWAS analysis

#### Fine-mapping

SusieR analysis allowed us to identify at least one credible set for 20 out of the 23 IPF locus signals and to fine-map a substantially reduced number of prioritised variants compared to the clumped signals with median credible set size of three (Supplementary Table 9).

Additionally, we mapped consequences to all variants belonging to each credible set and annotated the worst (i.e. predicted to be the most damaging) consequence across each set. This procedure enabled us to identify additional potential variants and genes that could underlie the associations (Supplementary Table 9).

We examined the novel signals around focal variants chr1:150579566_G_A and chr8:97822998_T_C and their credible sets in greater detail (Supplementary Figure *4*, Supplementary Figure *5*):

- The signal around focal variant chr1:150579566_G_A (rs16837903), was fine-mapped into a single credible set of 24 variants with lead credible set variant chr1:150598509_T_G; rs11204675; OR (95%CI) = 1.08 (1.05, 1.11; P-value=1.06x$10^{-7}$ (Supplementary Table 9). chr1:150598509_T_G does not present strong LD with the focal variant chr1:150579566_G_A (r2 < 0.4). The worst consequence across all variants within the identified credible set was presented by variant chr1:150575271_G_A (rs878471), which is a 3' UTR variant for the gene *MCL1* (Supplementary Table 9) and with a CADD score of 12.1 compared to the CADD score of 1.72 for chr1:150598509_T_G.
- The signal around focal variant chr8:97822998_T_C (rs6983263) was fine-mapped into a single credible set of 18 variants all found within gene LAPTM4B (Supplementary Figure 5). The focal variant was the lead credible set variant. The variant with the worst consequence within the credible set was chr8:97836681_A_T (rs10504980) located in an intron of *LAPTM4B* and has a CADD score of 8.9 compared to the CADD score of 0.04 for chr8:97822998_T_C (Supplementary Table 9).

*Supplementary Table 9. Fine-mapping results for lead variants from IPF meta-analysis and worst consequence variant in each credible set. Index CS indicates the credible set index for the focal variant fine-mapped window. nCS indicates the number of variants included in each credible set. Lead variant is the variant in the fine-mapped credible set with the lowest P-value. Consequence annotation for all variants across credible sets was generated using VEP v.105 and the worst consequence across transcripts was chosen. All variants were ranked according to their consequence type and CADD scores are provided for the lead variant and the variant with the worst consequence across all variants in each credible set.* ***Blue font*** *identifies novel locus signals of this study compared to Partanen et al. (2022).* ***Red font*** *identifies signals that did not successfully fine-map.*

| Locus | Focal CS variant | index CS | n CS | Lead CS variant | Lead CS Variant CADD | Worst CS variant | Worst CS CADD | Worst CS consequence | Worst CS gene |
|---|---|---|---|---|---|---|---|---|---|
| **1p36.23** | chr1:9107187_C_A | L1 | 14 | chr1:9107187_C_A | 1.95 | chr1:9102126:T:G | 7.9 | 3' UTR | *GPR157* |
| **1p31.1** | chr1:77998184_T_C | L1 | 14 | chr1:77998184_T_C | 4.51 | chr1:78062228:T:C | 7.0 | intron | *GIPC2* |
| **1q21.2** | **chr1:150579566_G_A** | **L1** | **24** | **chr1:150598509_T_G** | **1.72** | **chr1:150575271:G:A** | **12.1** | **3' UTR** | ***MCL1*** |
| **3p21.31** | chr3:44804157_T_C | L1 | 22 | chr3:44804157_T_C | 7.37 | chr3:44753389:A:C | 7.8 | missense | *KIAA1143* |
| **3q26.2** | chr3:169774313_C_T | L1 | 12 | chr3:169774313_C_T | 22.50 | chr3:169796797:A:T | 19.8 | missense | *LRRC34* |

| | | | | | | | | | |
|---|---|---|---|---|---|---|---|---|---|
| **4q22.1** | chr4:88915668_C_T | L1 | 3 | chr4:88915668_C_T | 0.25 | chr4:89113535:A:G | 7.8 | synonymous | *TIGD2* |
| **4q22.1** | chr4:88915668_C_T | L2 | 70 | chr4:88963935_G_T | 2.36 | chr4:88929112:A:G | 4.8 | intron | *FAM13A* |
| **5p15.33** | chr5:1282299_G_A | L1 | 1 | chr5:1279675_C_T | 1.36 | chr5:1287225:A:G | 3.2 | intron | *TERT* |
| **5p15.33** | chr5:1282299_G_A | L3 | 2 | chr5:1287079_G_A | 0.49 | chr5:1279675:C:T | 0.5 | intron | *TERT* |
| **5p15.33** | chr5:1282299_G_A | L2 | 1 | chr5:1442411_G_A | 0.23 | chr5:1442411:G:A | 0.2 | intron | *SLC6A3* |
| **5q35.1** | chr5:169477001_T_G | L1 | 1 | chr5:169477001_T_G | 5.03 | chr5:169477001:T:G | 5.0 | intergenic | . |
| **6p24.3** | chr6:7562999_T_G | L1 | 1 | chr6:7562999_T_G | 1.41 | chr6:7562999:T:G | 1.4 | intron | *DSP* |
| **6p21.33** | chr6:31380300_G_A | - | - | - | - | - | - | - | - |
| **7p22.3** | chr7:1937227_G_A | L1 | 91 | chr7:1937227_G_A | 5.09 | chr7:1936821:C:T | 7.8 | missense | *MAD1L1* |
| **7q22.1** | chr7:100007746_G_A | L1 | 4 | chr7:100007746_G_A | 0.01 | chr7:100031438:C:T | 2.7 | intron | *ZKSCAN1* |
| **7q32.1** | chr7:129095384_G_A | L1 | 88 | chr7:129095384_G_A | 2.43 | chr7:128948946:T:C | 3.2 | 3' UTR | *IRF5* |
| **8q22.1** | **chr8:97822998_T_C** | **L1** | **18** | **chr8:97822998_T_C** | **0.04** | **chr8:97836681:A:T** | **8.9** | **intron** | ***LAPTM4B*** |
| **9q31.3** | chr9:110842172_T_C | L1 | 5 | chr9:110842172_T_C | 3.81 | chr9:110842172:T:C | 3.8 | Regulatory region | . |
| **11p15.5** | chr11:1214934_A_G | L4 | 2 | chr11:1044640_C_T | 7.95 | chr11:1161648:G:A | 7.9 | intron | *MUC5AC* |
| **11p15.5** | chr11:1214934_A_G | L5 | 1 | chr11:1057948_G_C | 0.31 | chr11:1291288:C:T | 3.3 | intron | *TOLLIP* |
| **11p15.5** | chr11:1214934_A_G | L8 | 1 | chr11:1120870_T_G | 3.69 | chr11:1289967:G:T | 0.6 | intron | *TOLLIP* |
| **11p15.5** | chr11:1214934_A_G | L10 | 1 | chr11:1122434_TGGTCCTGACC_T | 1.34 | chr11:935428:A:ACC | 0.3 | intron | *AP2A2* |
| **11p15.5** | chr11:1214934_A_G | L7 | 1 | chr11:1161648_G_A | 0.02 | chr11:1288201:G:A | 0.02 | intron | *TOLLIP* |
| **11p15.5** | chr11:1214934_A_G | L1 | 1 | chr11:1214934_A_G | 7.26 | chr11:1214934:A:G | 7.3 | Upstream gene | *AC061979.1* |
| **11p15.5** | chr11:1214934_A_G | L3 | 1 | chr11:1216050_A_AC | 1.29 | chr11:1216050:A:AC | 1.3 | Upstream gene | *AC061979.1* |
| **11p15.5** | chr11:1214934_A_G | L2 | 2 | chr11:1224068_C_T | 0.60 | chr11:1057948:G:C | 0.6 | Downstream gene | *LINC02688* |
| **11p15.5** | chr11:1214934_A_G | L6 | 3 | chr11:1288201_G_A | 10.69 | chr11:1122434:TGGTCCTGACC:T | 10.7 | intergenic | . |
| **11p15.5** | chr11:1214934_A_G | L9 | 3 | chr11:1329907_A_T | 2.06 | chr11:1120870:T:G | 2.1 | intergenic | . |
| **13q34** | chr13:112881427_C_T | L1 | 3 | chr13:112881427_C_T | 0.22 | chr13:112886111:C:T | 4.6 | 3' UTR | *ATP11A* |
| **15q15.1** | chr15:40428343_G_T | - | - | - | - | - | - | - | - |
| **15q25.3** | chr15:85748038_C_T | L1 | 33 | chr15:85748038_C_T | 2.76 | chr15:85749240:T:A | 18.0 | 3' UTR | *AKAP13* |
| **16p13.3** | chr16:72352_C_T | L1 | 15 | chr16:72352_C_T | 5.44 | chr16:35561:C:T | 0.08 | non_coding transcript_exon | *IL9RP3* |
| **17q21.31** | chr17:46126154_C_T | - | - | - | - | - | - | - | - |
| **19p13.3** | chr19:4717660_A_G | L1 | 3 | chr19:4717660_A_G | 17.04 | chr19:4717660:A:G | 17.0 | missense | *DPP9* |
| **19p13.3** | chr19:4717660_A_G | L2 | 4 | chr19:5847989_T_C | 3.67 | chr19:5832198:G:A | 7.3 | missense | *FUT6* |
| **20q13.33** | chr20:63623343_A_G | L1 | 12 | chr20:63623343_A_G | 0.73 | chr20:63596004:G:A | 2.3 | intron | *GMEB2* |

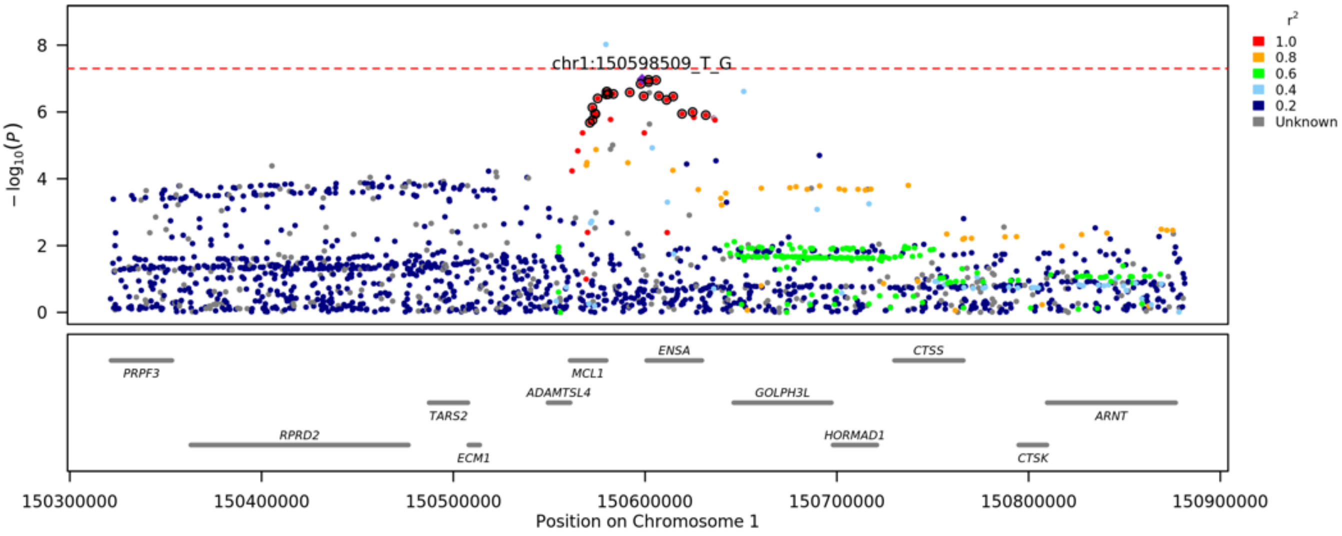

*Supplementary* Figure 4. *Fine-mapping Locuszoom-style plot for focal variant chr1:150579566_G_A using susieR. For this locus, a single credible set was identified with variant chr1:150598509_T_G having the lowest P-value in the credible set. The variants belonging to the credible set are represented as black outline circles. Gradient colours correspond to variant LD with chr1:150598509_T_G.*

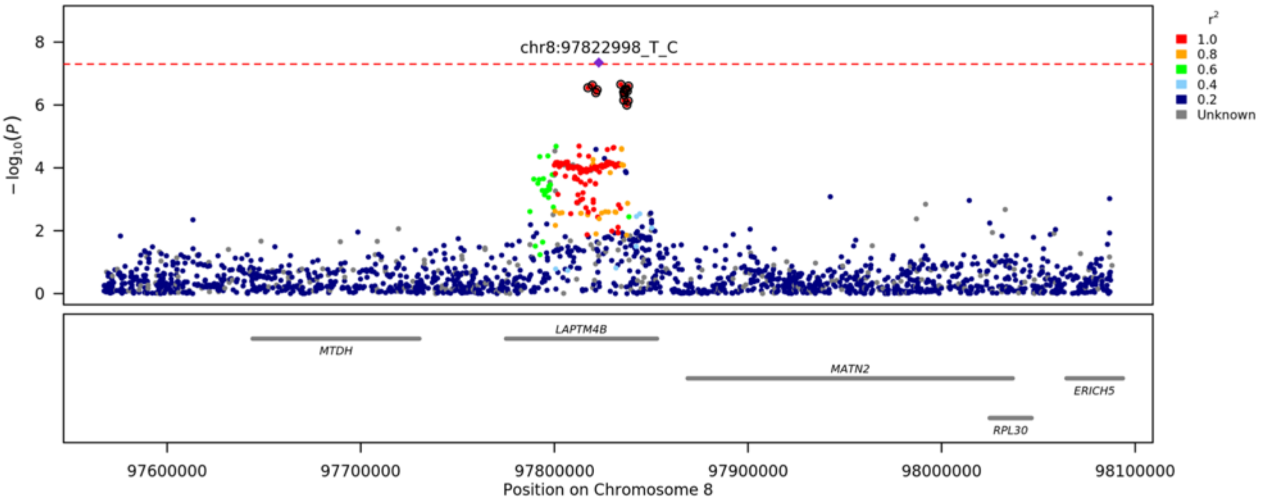

*Supplementary* Figure 5. *Fine-mapping Locuszoom-style plot for focal variant chr8:97822998_T_C using susieR. For this locus, a single credible set was identified with variant chr8:97822998_T_C having the lowest P-value in the credible set. The variants belonging to the credible set are represented as black outline circles. Gradient colours correspond to variant LD with chr8:97822998_T_C.*

### Aggregate variant burden testing (AVT) analysis

#### AVT Cohorts

*Supplementary Table 10. Cohorts used in AVT analysis*

| Cohort | Cases | Controls | Total | Case - Control Ratio | Mean Age (SD) | Percent Female |
|---|---|---|---|---|---|---|
| **IPF** | 569 | 50,847 | 51,416 | 1:89 | 42.4 (21.15) | 54.6% |

#### AVT variant Masks

*Supplementary Table 11. Four variant masks for AVT analysis. VEP impact scale for MODERATE includes: inframe insertion, inframe deletion, missense, protein altering, for HIGH includes transcript ablation, splice acceptor, splice donor, stop gained, frameshift, stop lost, start lost, transcript amplification. gnomAD allele frequencies are taken from v3.1.1 using the non-Finnish European (NFE) subset. Internal allele frequencies are taken from the IPF case-control cohort used in the specific analysis.*

| Description | gnomAD NFE AF Mask | Internal AF Mask | Variant Mask |
|---|---|---|---|
| **Loss-of-function (LOF)** | $\leq$0.1% | $\leq$0.1% | LOFTEE: High Confidence LoF |
| **Ultra-rare damaging (URD)** | $\leq$0.0005% | $\leq$0.025% | LOFTEE: High Confidence LoF **OR**<br>VEP Impact: MODERATE or HIGH with CADD >=20 |
| **Rare damaging (RD)** | $\leq$0.005% | $\leq$0.05% | LOFTEE: High Confidence LoF **OR**<br>VEP Impact: MODERATE or HIGH with CADD >=20 |
| **Flexible damaging (FD)** | LoF $\leq$0.1%<br>VEP MODERATE $\leq$ 0.5% | LoF $\leq$0.1%<br>VEP MODERATE $\leq$ 0.5% | LOFTEE: High Confidence LoF **OR**<br>VEP Impact: MODERATE or HIGH with CADD >=10 |

#### IPF cohort AVT results

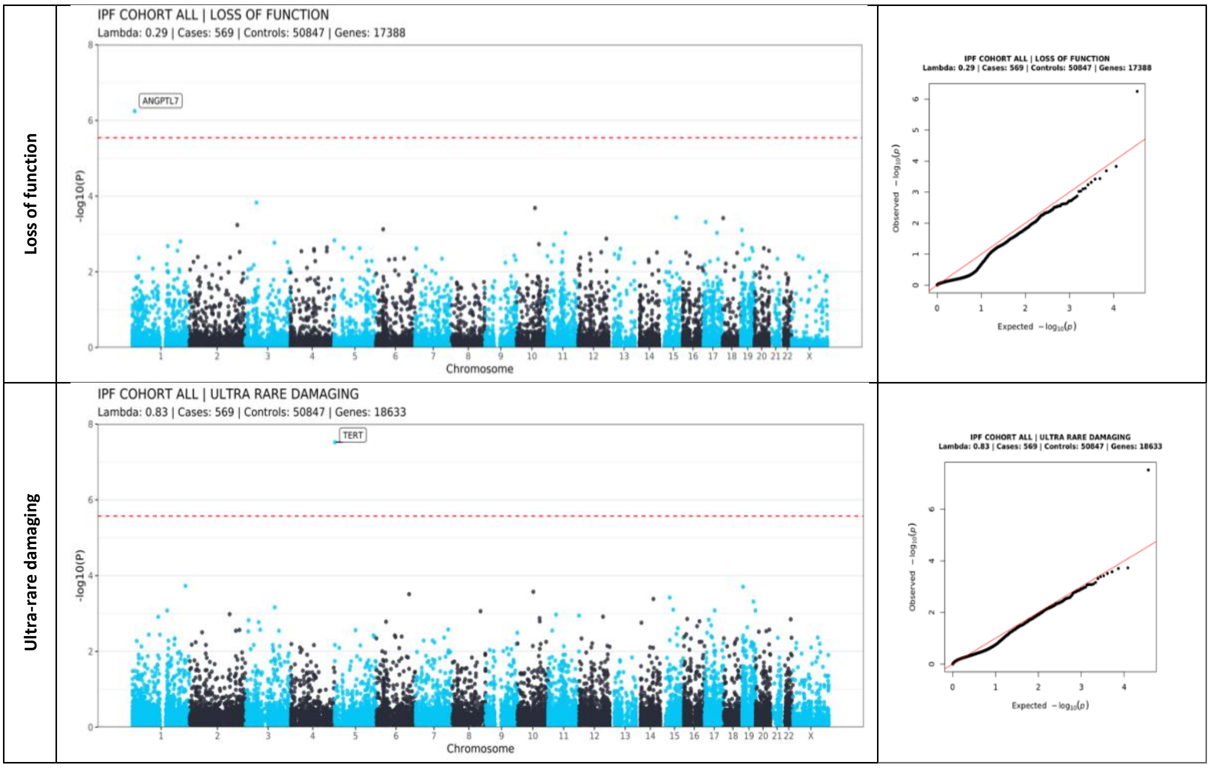

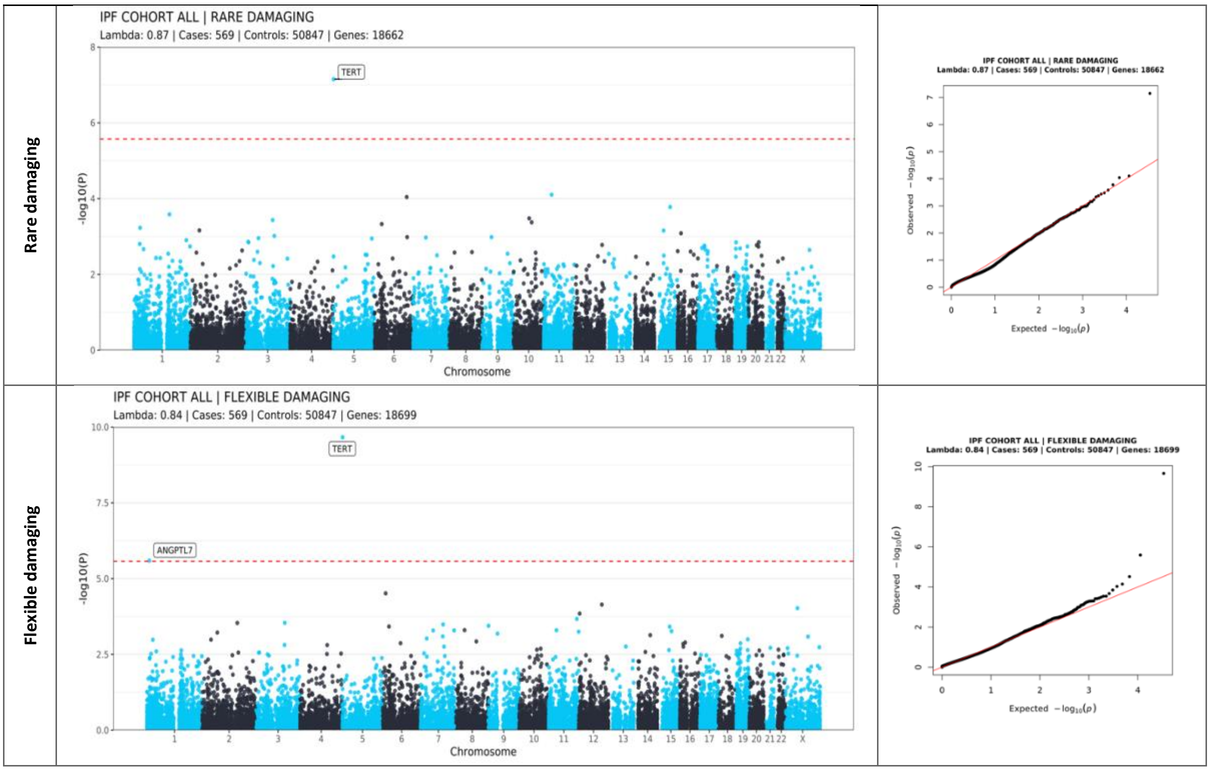

*Supplementary Figure 6: Manhattan and Q-Q plots for Aggregate Variant burden testing (AVT) analysis for the IPF cohort. Red dashed line indicates Bonferroni significance for within mask correction.*

#### AVT association statistics

*Supplementary Table 12. ANGPTL7 single variant association statistics (IPF cohort loss-of-function mask).*

| Variant Group | Variant ID | AC | AF | AF Cases | AF Controls | AF gnomAD 3.1.2 NFE* | OR (95% CI) | *P*-value |
|---|---|---|---|---|---|---|---|---|
| MAC≤10 (ultra_rare_variant) | chr1:11192318_CTG_C<br>chr1:11193665_G_A<br>chr1:11194575_C_CTCCAGTATCA<br>chr1:11195021_TA_T | 8 | 7.78E-05 | 0 | 7.87E-05 | Not assessed | 0.39 (0-99.76) | 0.739 |
| MAC>10 | chr1:11193631_C_T | 40 | 3.89E-04 | 7.91E-03 | 3.05E-04 | 3.24E-04 | 28.79 (8.51-97.43) | 6.73E-08 |

*NFE in gnomAD 3.1.2 corresponds to non-Finnish Europeans.

*Supplementary Table 13. ANGPTL7 single variant association statistics from coding AVT (output of SAIGE-GENE+).*

| Mask | Marker ID | AC | AF | AF Cases | AF Controls | AF gnomAD 3.1.2 NFE | BETA (Standard Error) | *P*-value |
|---|---|---|---|---|---|---|---|---|
| LOF | ultra_rare_variant (4) | 8 | 7.78E-05 | 0 | 7.87E-05 | Not assessed | -0.944 (2.83) | 0.739 |
| | chr1:11193631_C_T | 40 | 3.89E-04 | 7.91E-03 | 3.05E-04 | 3.24E-04 | 3.36 (0.622) | **6.73E-08** |
| FD | ultra_rare_variant (65) | 158 | 1.54E-03 | 8.79E-04 | 1.54E-03 | Not assessed | -0.352 (0.856) | 0.681 |
| | chr1:11189667_T_A | 33 | 3.21E-04 | 8.79E-04 | 3.15E-04 | 3.97E-04 | 1.77 (1.68) | 0.294 |
| | chr1:11189732_G_C | 33 | 3.21E-04 | 0 | 3.25E-04 | 3.97E-04 | -1.03 (2.51) | 0.68 |
| | chr1:11189746_T_C | 41 | 3.99E-04 | 0 | 4.03E-04 | 2.50E-04 | -1.07 (1.57) | 0.497 |
| | chr1:11192300_A_G | 53 | 5.15E-04 | 0 | 5.21E-04 | 5.73E-04 | -1.05 (1.59) | 0.508 |
| | chr1:11192312_G_A | 283 | 2.75E-03 | 8.84E-04 | 2.77E-03 | 2.75E-03 | -0.664 (0.641) | 0.3 |
| | chr1:11193631_C_T | 40 | 3.89E-04 | 7.91E-03 | 3.05E-04 | 3.24E-04 | 3.85 (0.711) | **6.28E-08** |

| | | | | | | | | |
|---|---|---|---|---|---|---|---|---|
| | chr1:11193659_G_A | 13 | 1.26E-04 | 0 | 1.28E-04 | 5.88E-05 | -1.05 (2.94) | 0.72 |
| | chr1:11193761_G_A | 11 | 1.07E-04 | 0 | 1.08E-04 | 2.94E-05 | -1.03 (3.86) | 0.79 |
| | chr1:11194982_C_T | 10 | 9.72E-05 | 0 | 9.83E-05 | 4.41E-05 | -1.02 (4.2) | 0.807 |

*LOF: loss-of-function, FD: flexible damaging. AC: Allele Count, AF: Allele Frequency. Variants with MAC≤10 are grouped into an ultra_rare_variant. The brackets indicate the number of variants of MAC≤10 in the ultra_rare_variant group. Note that effect size estimates and P-values for the same variant included in different masks can differ slightly as they are derived from saddle point approximation. NFE in gnomAD 3.1.2 corresponds to non-Finnish Europeans.*

#### Transcriptome-wide association study (TWAS)

*Supplementary Table 14. Results from Transcriptome-wide association study (TWAS) for IPF using S-prediXcan and GTEX v8 expression data from lung tissue. S-PrediXcan's association results (zscore and P-value) for each gene, the number of snps in the model (n_snps) and success indicator for colocalization (iColoc) check are shown. Results for genes with P-value < $3.44x10^{-6}$ (Bonferonni adjusted threshold). Ref displays the reference where gene was first reported from TWAS analysis with star (*) indicating the present study.*

| Gene name | zscore | *P*-value | n_snps | iColoc | Ref |
|---|---|---|---|---|---|
| ***MUC5B*** | 23.61 | 3.35E-123 | 1 | yes | [2] |
| ***DSP*** | -13.71 | 9.00E-43 | 1 | yes | [2] |
| ***BAHD1*** | -9.80 | 1.16E-22 | 2 | no | [2] |
| ***MAPT*** | -9.36 | 8.18E-21 | 1 | yes | [2] |
| ***ACTRT3*** | -9.10 | 8.97E-20 | 2 | no | * |
| ***ARHGAP27*** | -7.91 | 2.51E-15 | 1 | no | [3] |
| ***WNT3*** | -7.81 | 5.52E-15 | 2 | no | [2] |
| ***MYNN*** | 7.73 | 1.09E-14 | 2 | yes | [3] |
| ***KIF15*** | -6.61 | 3.91E-11 | 1 | no | * |
| ***TOLLIP*** | 6.11 | 1.03E-09 | 2 | no | [2] |
| ***LRRC37A*** | -6.09 | 1.13E-09 | 4 | no | [2] |
| ***CD151*** | -6.00 | 2.02E-09 | 4 | no | * |
| ***BRSK2*** | 5.75 | 9.05E-09 | 1 | no | [2] |
| ***TNPO3*** | 5.62 | 1.91E-08 | 2 | yes | * |
| ***AP2A2*** | 5.41 | 6.37E-08 | 2 | no | * |
| ***TRIM10*** | -5.20 | 2.02E-07 | 2 | no | * |
| ***SERTAD2*** | 5.13 | 2.96E-07 | 1 | yes | * |
| ***FLOT1*** | 4.95 | 7.34E-07 | 3 | no | * |
| ***ZSCAN16*** | -4.93 | 8.40E-07 | 1 | no | * |
| ***RMDN3*** | -4.83 | 1.36E-06 | 1 | no | * |
| ***DEPTOR*** | -4.66 | 3.13E-06 | 2 | yes | [2] |

#### IPF and COVID-19

##### IPF and COVID-19 summary statistics

*Supplementary Table 15. Case-control breakdown for IPF and severe COVID-19 summaries used for colocalisation analysis.*

| Trait | Cohort (study) | Cases | Controls | Type |
|---|---|---|---|---|

| | | | | |
|---|---|---|---|---|
| **IPF** | GBMI + Allen et al. + 100kGP (This study) | 11,746 | 1,416,493 | GWAS meta-analysis |
| **Severe COVID-19** | COVID-19 Host Genetics Initiative freeze 7 (HGI V7, B2_ALL_leave_23andme) | 44,986 | 2,356,386 | GWAS meta-analysis |

#### IPF and COVID-19 colocalisation

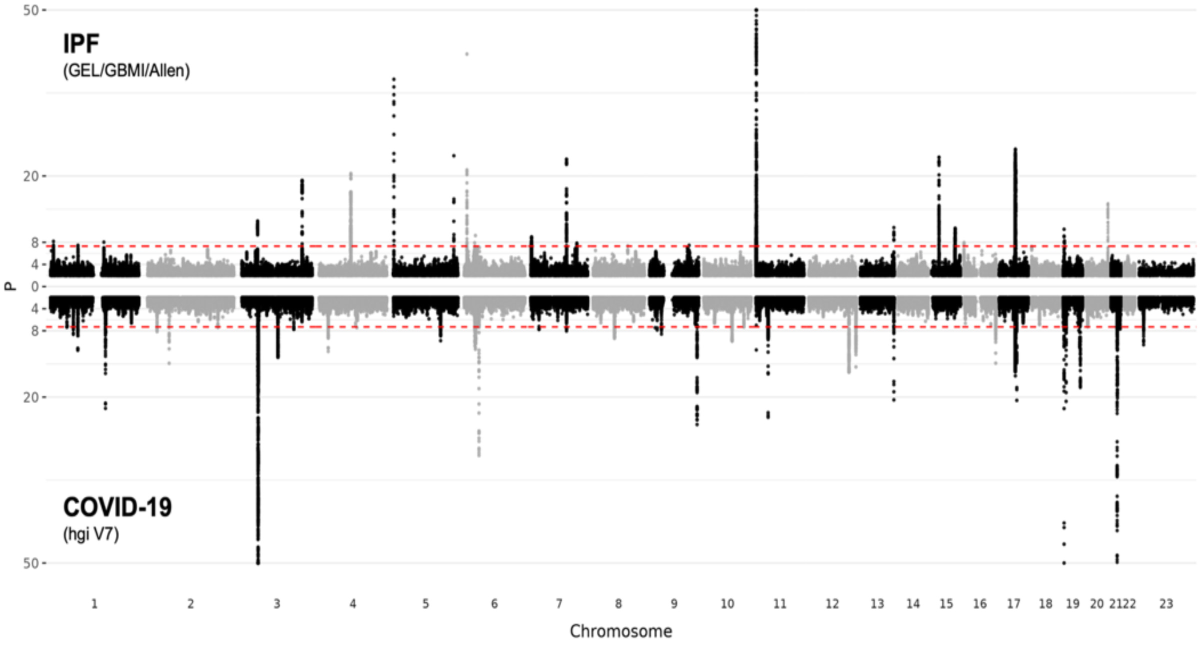

*Supplementary Figure 7. Miami plot of IPF and severe COVID GWAS. Red dashed line corresponds to P-value < x10-8. Red dashed line corresponds to P-value < 5x10$^{-8}$. P-values below 10$^{-50}$, are set to 10$^{-50}$.*

#### IPF and COVID-19 genetic correlation

To calculate the genetic correlation ($r_g$) between IPF and severe COVID-19, we used an LD-score regression-based approach as implemented in LDSC (Methods). The estimated $r_g$ was 0.39 and significantly different than zero (95%CI: 0.25, 0.53; *P*-value: 6.12x10$^{-8}$; Supplementary Table 16). We obtained a higher estimate for $r_g$ than the most recent previous estimate[5], with tighter confidence intervals and a statistically stronger *P*-value, due to the larger sample sizes used in this study (Supplementary Table 16). Our results further strengthen the support for a moderate correlation between IPF and severe COVID-19.

*Supplementary Table 16. Genetic correlation estimates in Partanen et al. (2022) and this study.*

| **Study** | **IPF** | | **Severe COVID-19** | | **$r_g$ [95% CI]** | ***P*-value** |
|---|---|---|---|---|---|---|
| | N cases | N controls | N cases | N controls | | |
| Partanen et al. 2022[5] | 11,160 | 1,364,410 | 24,274 | 2,061,529 | 0.31 [0.15, 0.47] | 1.0x10$^{-4}$ |
| This study | 11,746 | 1,416,493 | 44,986 | 2,356,386 | 0.39 [0.25, 0.53] | 6.1x10$^{-8}$ |

### Gene functional follow ups

#### MCL1 tissue-specific expression profile

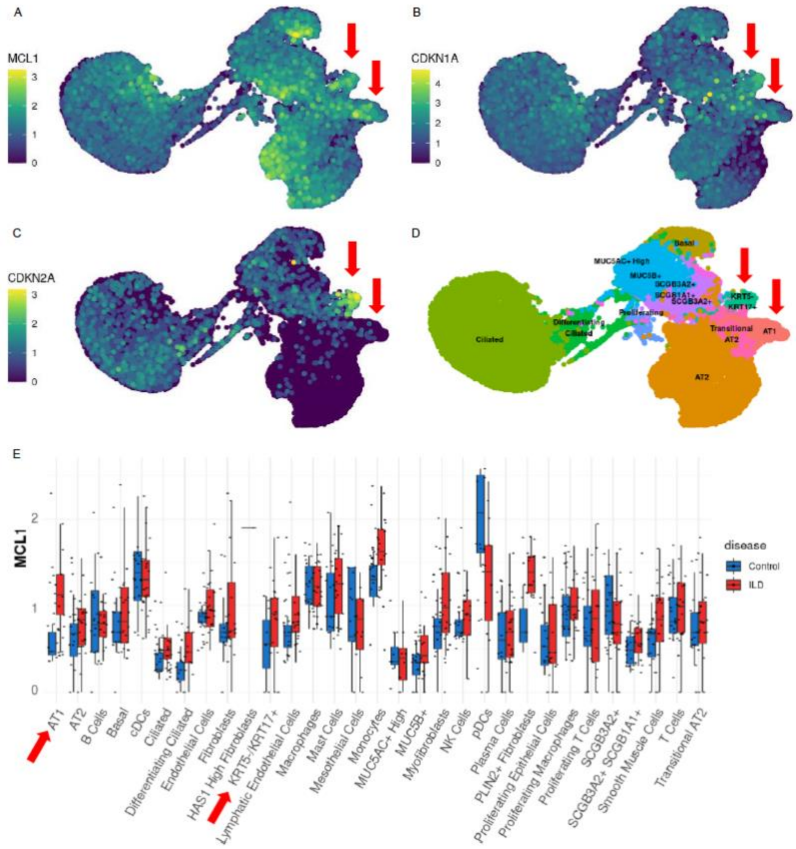

*Supplementary* Figure 8. Expression profile for *MCL1* for lung tissues *(A) MCL1 expression in epithelial cells. (B) CDKN1A expression in epithelial cells. (C) CDKN1A expression in epithelial cells. (D) Cell type annotation. (E) MCL1 expression across the whole dataset, averaged by patient, split by disease. All data from Banovich/Kropski dataset (patients n=30; GSE135893). Taken from IPF Cell Atlas epithelial cell experiments.*

#### ANGPTL7 potential functional significance for IPF

ANGPTL7 is part of a functionally heterogeneous family of angiopoietin-like proteins (ANGPTLs), comprising ANGPTL1 through to ANGPTL8. ANGPTLs are widely expressed in multiple tissues and have been implicated in diverse processes including angiogenesis, cancer, inflammation, metabolism, and haematopoiesis (reviewed in Carbone et al. 2018[6]). The precise function of ANGPTLs has been difficult to define due to their pleiotropic and sometimes context-dependent activities. In addition, the relevant signalling pathways are incompletely understood, although several receptors and downstream transcriptional effectors have been identified[7–9].

ANGPTL7 is one of the less well characterised members of the family and its receptor remains to be identified. The following highlights three contexts in which ANGPTL7 function has been studied:

- Multiple functional and genetic studies have linked *ANGPTL7* to glaucoma. Specifically, two independent studies [10,11] have found that the aforementioned Arg177Ter (rs143435072) variant and the rare missense variant Gln175His (rs28991009) provide protection against glaucoma through a reduction in intraocular pressure (IOP). ANGPTL7 is expressed in the trabecular meshwork (TM), an ECM-rich structure that regulates IOP. ANGPTL7 activity is thought to promote ECM formation in the TM, thereby inhibiting the outflow of aqueous humour and raising intraocular pressure[12]. The molecular mechanism remains poorly understood but may involve ANGPTL7 inducing the expression of ECM components through an as-yet unidentified receptor.

- In the eye, ANGPTL7 is also expressed in corneal keratocytes. Mouse model and cell culture experiments suggest that ANGPTL7 has anti-angiogenic activity and that this is required for maintaining the avascularity of the cornea[13].
- ANGPTL7 has been reported to be overexpressed in tumour samples and tumour cell lines[14], consistent with what has been reported for other ANGPTLs (reviewed in Carbone et al. 2018[6]). Mouse model and cell culture experiments suggest that ANGPTL7 has pro-angiogenic activity on differentiated endothelial cells, thereby driving tumour growth. Whilst this appears to contradict findings in the cornea, evidence of anti-angiogenic activity in one tissue and pro-angiogenic in another has also been reported for ANGPTL4[15,16].

To gain insight on the potential role of *ANGPTL7* in the lung, a review was performed of the expression of *ANGPTL7* in the lung using IPF Cell Atlas, a database of single-cell differential expression studies carried out in fibrotic and healthy control lungs[17].

Amongst epithelial cell types, *ANGPTL7* is sparsely but specifically expressed in type-1 alveolar epithelial cells (AT1) across four independent single-cell expression datasets (Supplementary Figure 9). This expression pattern is specific to *ANGPTL7* (Supplementary Figure 10), as other ANGPTLs are either not expressed in AT1 cells (ANGPTLs 3, 5, 6, and 8) or are expressed more broadly across different cell types in the lung (ANGPTLs 1, 2, and 4). Amongst mesenchymal cell types, *ANGPTL7* is strongly expressed in lung fibroblasts (Supplementary Figure 11).

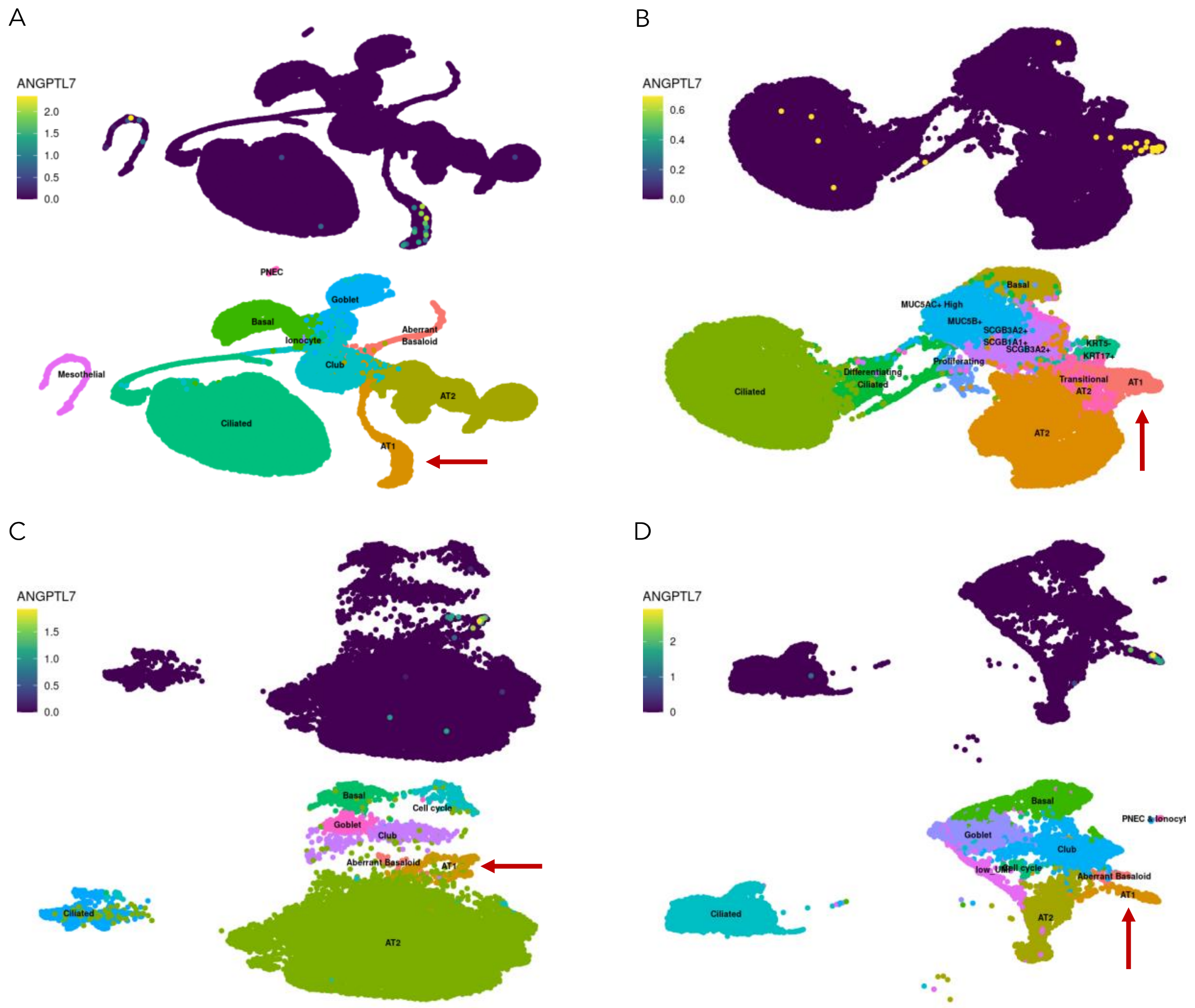

*Supplementary Figure 9. ANGPTL7 is sparsely but specifically expressed in type-1 cells. Taken from IPF Cell Atlas epithelial cell experiments. (A) Kaminski/Rosas, n=79 (B) Banovich/Kropski, n=30, (C) Misharin, n=16, (D) Lafyatis n=6. Arrow indicates AT1 population.*

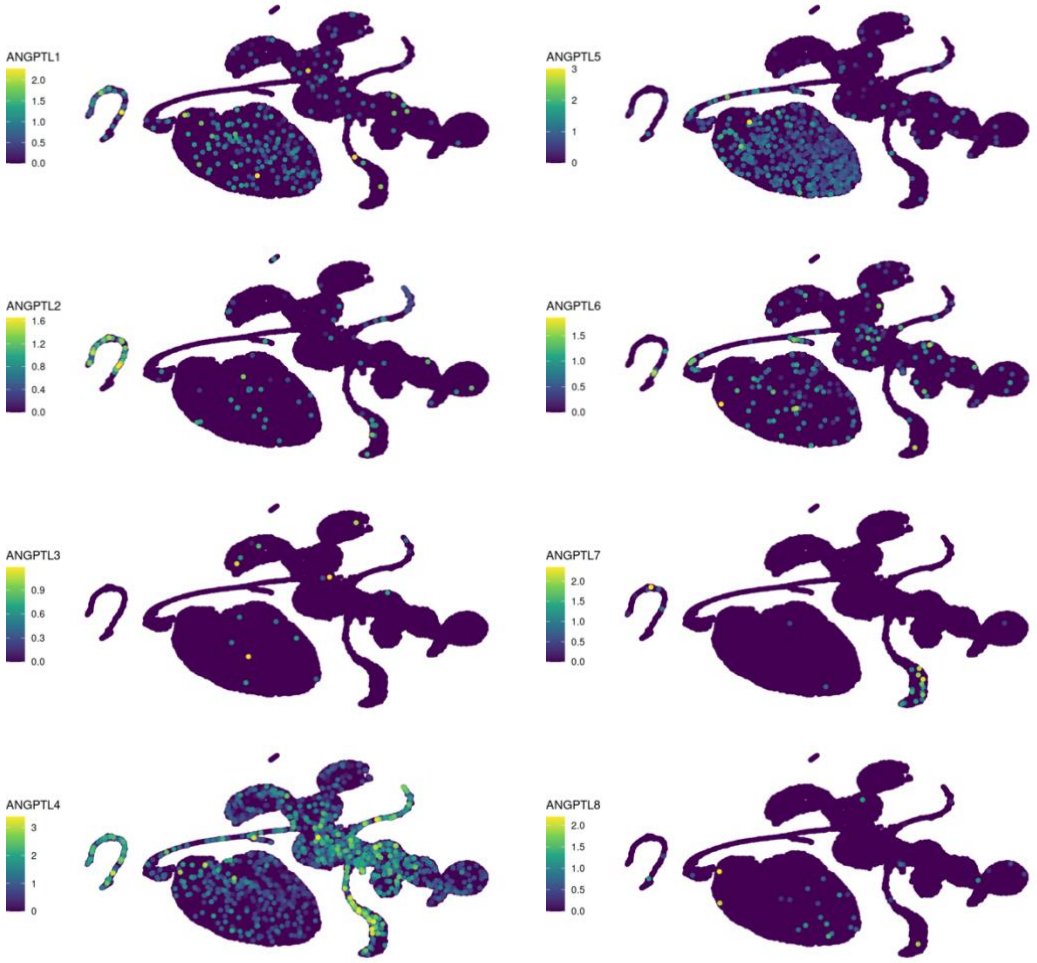

*Supplementary Figure 10. Narrow expression in type-1 cells is specific to ANGPTL7. Taken from IPF Cell Atlas cell experiments from Kaminski/Rosas, n=79.*

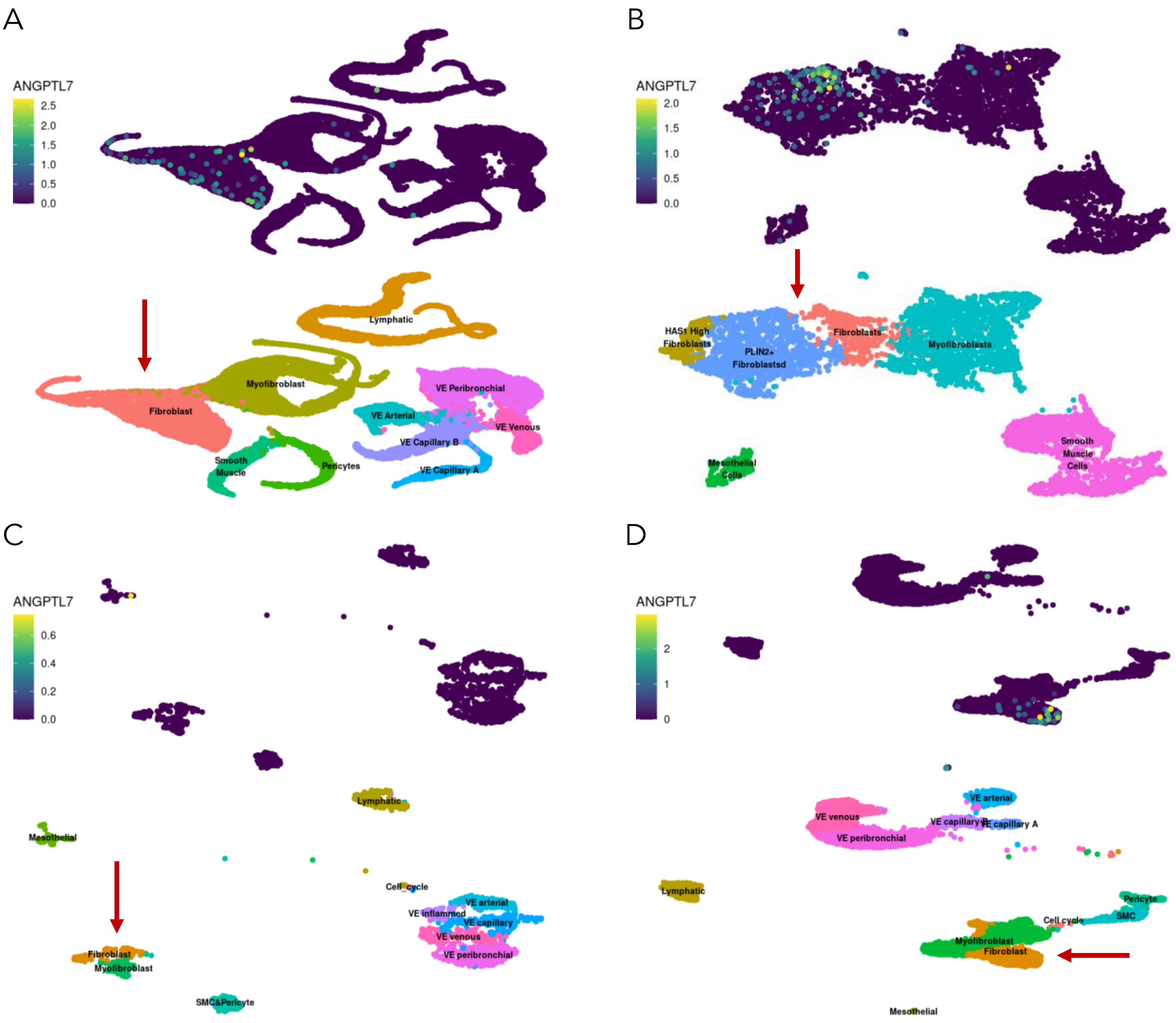

*Supplementary Figure 11: ANGPTL7 is expressed in fibroblasts. Taken from IPF Cell Atlas epithelial cell experiments. (A) Kaminski/Rosas, n=79 (B) Banovich/Kropski, n=30, (C) Misharin, n=16, (D) Lafyatis n=6. Arrow indicates fibroblast population (PLIN2+ fibroblasts in Banovich/Kropski).*

The differences in *ANGPTL7* expression between healthy and fibrotic lungs in the IPF Cell Atlas were reviewed. Whilst *ANGPTL7* appears to be differentially expressed in some of the datasets, the effect is not consistent across all datasets. In the Kaminski/Rosas and the Lafyatis datasets, *ANGPTL7* expression is absent in type-1 cells IPF lungs. In the Misharin dataset, *ANGPTL7* expression in type-1 cells is reduced in lungs of patients with interstitial lung disease (ILD). However, *ANGPTL7* expression in type-1 cells is slightly elevated in ILD lungs in the Banovich/Kropski dataset. It deserves further investigation whether the lack of consistency could be attributable to differences in biopsy protocol or tissue representation. Loss or reduction of *ANGPTL7* expression in type-1 cells for IPF patients is supportive of the genetic results for IPF risk increase due to disruption/reduction in *ANGPTL7* expression from missense mutations.

The alveolar epithelium comprises type-1 and type-2 epithelial cells. Type-1 cells line 95% of the alveolar surface, enabling gas exchange by virtue of their squamous morphology (reviewed in Wang et al. 2020[18]). Type-2 cells are progenitor cells that regenerate type-1 cells in response to lung injury and as part of natural tissue turnover. Existing pulmonary fibrosis disease genes are thought to act in type-2 cells (reviewed in Parimon et al. 2019[19]). For example, rare variants in *SFTPA1*, *SFTPA2*, and *SFTPC* disrupt secretion of pulmonary surfactant by type-2 cells. Rare and common variants in telomere maintenance genes (e.g., *TERT*, *TERC*, *RTEL1*, *PARN*) are thought to promote pulmonary fibrosis by reducing the replicative potential of type-2 cells, thereby interfering with their ability to regenerate type-1 cells. *ANGPTL7* is the first candidate disease gene that is expressed specifically in type-1 cells. Therefore, *ANGPTL7* could have considerable translational interest, as damage to type-1 cells and their replacement by fibrotic tissue directly drives alveolar fibrosis.

Given the above, we speculate on the disease mechanism of *ANGPTL7* in pulmonary fibrosis:

- *ANGPTL7* may act as an autocrine or paracrine signal secreted by type-1 cells and/or lung fibroblasts to promote interstitial ECM formation, analogous to its role in the TM of the eye. Loss of *ANGPTL7* would disrupt the alveolar ECM, which would either render the alveolar epithelium more susceptible to damage or directly promote fibroblast activation. This would be consistent with evidence suggesting that ECM abnormalities are a cause (rather than merely a consequence) of pulmonary fibrosis[20,21].
- *ANGPTL7* may be secreted by type-1 cells and/or lung fibroblasts to inhibit alveolar angiogenesis, analogous to its putative role in the cornea. Loss of *ANGPTL7* would lead to excess alveolar vascularisation, thereby promoting fibrosis. Consistent with this hypothesis, hepatic angiogenesis is closely linked to progression of fibrosis in chronic liver disease[22]. In addition, vascular abnormalities are one of the histological hallmarks of IPF. However, it has not been demonstrated that these abnormalities are a cause rather than a consequence of pulmonary fibrosis, and the importance of vascularisation as a driver of pulmonary fibrosis remains unclear[23].

Testing the above hypotheses and elucidating the molecular and cellular role of *ANGPTL7* in the lung will likely require loss- and gain-of-function experiments in cell-based assays and in mouse models.

#### References

1. Partanen, J. J. *et al.* Leveraging global multi-ancestry meta-analysis in the study of idiopathic pulmonary fibrosis genetics. (2022) doi:10.1016/j.xgen.2022.100181.
2. Gong, W., Guo, P., Liu, L., Guan, Q. & Yuan, Z. Integrative Analysis of Transcriptome-Wide Association Study and mRNA Expression Profiles Identifies Candidate Genes Associated With Idiopathic Pulmonary Fibrosis. *Front Genet* **11**, (2020).
3. Chen, M. *et al.* Integrative analyses for the identification of idiopathic pulmonary fibrosis-associated genes and shared loci with other diseases. *Thorax* **78**, (2023).
4. Allen, R. J. *et al.* Genetic overlap between idiopathic pulmonary fibrosis and COVID−19. *European Respiratory Journal* **60**, (2022).
5. Partanen, J. J. *et al.* Leveraging global multi-ancestry meta-analysis in the study of idiopathic pulmonary fibrosis genetics. *Cell Genomics* **2**, 100181 (2022).
6. Carbone, C. *et al.* Angiopoietin-Like Proteins in Angiogenesis, Inflammation and Cancer. *Int J Mol Sci* **19**, (2018).
7. Camenisch, G. *et al.* ANGPTL3 stimulates endothelial cell adhesion and migration via integrin alpha vbeta 3 and induces blood vessel formation in vivo. *J Biol Chem* **277**, 17281–17290 (2002).
8. Zheng, J. *et al.* Inhibitory receptors bind ANGPTLs and support blood stem cells and leukaemia development. *Nature* **485**, 656–660 (2012).
9. Ben-Zvi, D. *et al.* Angptl4 links α-cell proliferation following glucagon receptor inhibition with adipose tissue triglyceride metabolism. *Proc Natl Acad Sci U S A* **112**, 15498–15503 (2015).
10. Praveen, K. *et al.* ANGPTL7, a therapeutic target for increased intraocular pressure and glaucoma. *Commun Biol* **5**, 1051 (2022).
11. Tanigawa, Y. *et al.* Rare protein-altering variants in ANGPTL7 lower intraocular pressure and protect against glaucoma. *PLoS Genet* **16**, (2020).
12. Comes, N., Buie, L. K. K. & Borrás, T. Evidence for a role of angiopoietin-like 7 (ANGPTL7) in extracellular matrix formation of the human trabecular meshwork: implications for glaucoma. *Genes Cells* **16**, 243–259 (2011).
13. Toyono, T. *et al.* Angiopoietin-like 7 is an anti-angiogenic protein required to prevent vascularization of the cornea. *PLoS One* **10**, (2015).
14. Parri, M. *et al.* Angiopoietin-like 7, a novel pro-angiogenetic factor over-expressed in cancer. *Angiogenesis* **17**, 881–896 (2014).
15. Le Jan, S. *et al.* Angiopoietin-like 4 is a proangiogenic factor produced during ischemia and in conventional renal cell carcinoma. *Am J Pathol* **162**, 1521–1528 (2003).
16. Okochi-Takada, E. *et al.* ANGPTL4 is a secreted tumor suppressor that inhibits angiogenesis. *Oncogene* **33**, 2273–2278 (2014).
17. Neumark, N., Cosme, C., Rose, K. A. & Kaminski, N. The Idiopathic Pulmonary Fibrosis Cell Atlas. *Am J Physiol Lung Cell Mol Physiol* **319**, L887–L893 (2020).
18. Wang, Y. *et al.* Pulmonary alveolar type I cell population consists of two distinct subtypes that differ in cell fate. *Proc Natl Acad Sci U S A* **115**, 2407–2412 (2018).
19. Parimon, T., Yao, C., Stripp, B. R., Noble, P. W. & Chen, P. Alveolar Epithelial Type II Cells as Drivers of Lung Fibrosis in Idiopathic Pulmonary Fibrosis. *Int J Mol Sci* **21**, (2020).
20. Parker, M. W. *et al.* Fibrotic extracellular matrix activates a profibrotic positive feedback loop. *J Clin Invest* **124**, 1622–1635 (2014).

21. Burgess, J. K., Mauad, T., Tjin, G., Karlsson, J. C. & Westergren-Thorsson, G. The extracellular matrix - the under-recognized element in lung disease? *J Pathol* **240**, 397–409 (2016).
22. Bocca, C., Novo, E., Miglietta, A. & Parola, M. Angiogenesis and Fibrogenesis in Chronic Liver Diseases. *Cell Mol Gastroenterol Hepatol* **1**, 477–488 (2015).
23. Hanumegowda, C., Farkas, L. & Kolb, M. Angiogenesis in pulmonary fibrosis: too much or not enough? *Chest* **142**, 200–207 (2012).
